# Early detection of seasonality and second-wave prediction in the COVID-19 pandemic

**DOI:** 10.1101/2020.09.02.20187203

**Authors:** Márcio Watanabe

## Abstract

Seasonality plays an essential role in the dynamics of many infectious diseases. Its confirmation in an emerging infectious disease is usually done using time series data from several years. By using statistical regression methods for time-series data pooled from more than 50 countries from both hemispheres, we show how to determine its presence in a pandemic at the onset of the seasonal period. We measure its expected effect in the mean transmission rate of SARS-coV-2 and predict when further epidemic outbreaks of COVID-19 will occur. The obtained result in the Northern Hemisphere shows that seasonality reduced the mean growth rate in 222.5% in April 2020. A relative reduction greater than 100% should be interpreted as a reduction changing an increasing rate to a decreasing one. In contrast, at the same moment, the seasonal effect in the Southern Hemisphere increased the mean growth rate in 740.3%. Our analysis simultaneously considers other confounding factors to properly separate them from seasonal effects and, in addition, we measure the mean global effect of social-distancing interventions and its relation with income. Future COVID-19 waves are expected to occur in autumn/winter seasons, typically between September and March in the Northern Hemisphere, and between April and September in the Southern Hemisphere. Simulations of a seasonal SEIR model with a social distancing effect are shown to describe the behavior of COVID-19 outbreaks in several countries. These results provide vital information for policy makers to plan their actions against the new coronavirus disease, particularly in the optimization of social-distancing interventions and vaccination schedules. Ultimately, our methods can be used to identify and measure seasonal effects in a future pandemic.

## 1. Introduction

During the COVID-19 pandemic, many public authorities made decisions based on predictions drawn from epidemiological compartmental models. The most prominent, and one of the simplest, of these models is the classical SEIR model. It can be seen as a qualitative epidemic model, since it is helpful in understanding the qualitative behavior of the dynamics of an epidemic. However, the use of such a simple model to make quantitative predictions mainly for long-term variables, such as the total size of epidemics, seems like an oversimplification.^1,2^

The COVID-19 pandemic is a complex phenomenon in which many other factors must be taken into consideration in order to obtain both qualitative understanding and quantitative predictive power. While the exponential growth of a SEIR model is better suited to model a single epidemic in a homogeneous, ^2^ closed system like a small town, more sophisticated models, such as meta-population models or agent-based models^3^, are more appropriate in the dynamics of interconnected, open systems with several cities and countries, as in a pandemic. The existence of several subgroups with considerably different epidemiological characteristics, such as mobility, makes the homogeneous assumption obsolete.^4^ The social distancing measures used to decrease the transmission rates around the world added more difficulty to these predictive models. Although several factors appear to influence the dynamics of the COVID-19 pandemic, there is an essential, well-known epidemiological phenomenon that has so far been absent in most of the models used by scientists and health officials: COVID-19 seasonality.

At some level, many infectious diseases, particularly viral respiratory ones, have a seasonal pattern of transmission.^5,6^ This implies that there is a period of the year in which the transmission rate is the highest and major epidemics are observed, as opposed to a complementary period of the year in which the transmission rate is significantly lower.

Influenza viruses, pneumonia, rotavirus, cholera, measles, dengue and other coronaviruses are some of the many infectious diseases in which seasonality plays an important role in the transmission rate.^5-13^

It is important to note that seasonality should not be confused with temperature. Although temperature is an important factor that is positively correlated with seasonality, there are many other factors that also influence this complex phenomenon. Climate factors, host behavior factors and biological factors, among others, can be associated with seasonal forces, such as precipitation, latitude, human mobility, school calendar, immunity and many others.^1,5,8,9^ In addition, these factors vary from place to place and must therefore be put in a relative perspective. For example, in tropical areas, an average temperature of 15 degrees Celsius is associated with winter days, whereas in higher latitudes it is associated with summer days. Hence, careful analysis is necessary when quantifying the relationship between temperature and seasonal transmission force and data from various locations.^7^ Therefore, seasonal forces can be more clearly measured by comparing data from Northern and Southern Hemispheres, since pooling data provides a reduction in data variability.

Respiratory syndromes have a common pattern in which the transmission rate is typically higher in autumn and winter and lower in spring and summer.^6,7^ Therefore, it is worth noting that the months with the highest number of cases are reversed in the Northern and Southern Hemispheres.

When considering the seasonal forces in modeling the dynamics of the disease, the β transmission rate will usually be given not by a constant positive real number, as in the basic SEIR model, but by a non-constant function of time β_t_. For clarity, consider that β_t_ assumes only two values β_max_ and β_min_, where β_max_ > β_min_ > 0. That is, we divide the year into two periods: one in which the transmission force is greater, with a transmission rate β_max_, and another in which the transmission force is lower, with β_min_ transmission rate. We can also associate other epidemiological parameters with β_max_ and β_min_, such as the basic reproduction number R0. If R0 is measured during a high seasonal period where β(t) = β_max_, then we will obtain R0 = R0_max_. However, if R0 is measured outside the seasonal period, β(t) = β_min_, and we will obtain R0=R0_min_, where R0_max_ > R0_min_.

Therefore, seasonality is fundamental in the dynamics of seasonal diseases and also in long-term forecasts.^1^ In particular, second wave forecasts are notably influenced by seasonality. An early detection of seasonality in an emergent disease is crucial for the public health authorities in planning and preventing a seasonal disease.^1^ The World health organization officially declared COVID-19 a pandemic at March 11. In this article, we show that seasonality of COVID-19 could have been detected less than two months after this date, using reported data of the pandemic from March and April, we give consistent statistical evidence that seasonal forces drives Sars-Cov2 transmission and discuss the important consequences which derive from it.

## 2. Results

Below, we show that COVID-19 transmission is highly affected by seasonality and how to identify the seasonality of an emerging pandemic disease. Using pooled data from the 50 largest outbreaks by the end of April 2020, we showed that there was already enough statistical evidence with less than two months of pandemic to conclude that the transmission of SARS-Cov2 is strongly influenced by seasonality. Normally, the seasonality of a disease would be confirmed using data from several years. Early detection of seasonality was possible because we use data from several simultaneous outbreaks, properly compared with data from the H1N1 2009 pandemic, which produces enough statistical information to attest to the effect of seasonality in a period of change in its transmission rate, the onset of a high seasonal period.

The analysis of the time series epidemiological data has been of great importance in COVID19 pandemic.^16,17^ Pooled methods decrease individual variability, increase statistical power providing timely analysis crucial in a period of urgency period such as a pandemic.^16^

To avoid confusion, have also calculated social distancing effects and measure their interactions through a multiple linear regression analysis together with a social-economic factor. In the supplementary appendix, we describe the simulation results of a seasonal SEIR model with temporary social distancing effect and compare its possible qualitative different scenarios with data from COVID-19 outbreaks of several countries. This analysis confirms that seasonality and social distancing interventions are fundamental to understanding the complexity of the COVID-19 epidemic curves.

We initially estimated that the global COVID-19 mean seasonal period coincides with the mean seasonal period of other respiratory syndromes, particularly with the mean seasonal period of H1N1 2009 pandemic, which occurred from September to March in the Northern Hemisphere, and from April to September in the Southern Hemisphere^10^. In these seasonal periods, the transmission rate is higher and larger epidemics are expected. Formally, we call the time interval where β = β_max_ as the high seasonal period.

However, there is some natural variability in data of endemic diseases. Thus, we also use data of another pandemic in order to find epidemiological data from several countries in a synchronized way. Figure 1 presents data from the 2009/2010 H1N1 pandemic in the Northern and Southern Hemispheres, taken from the World Health Organization database.^15^

**Figure 1.**
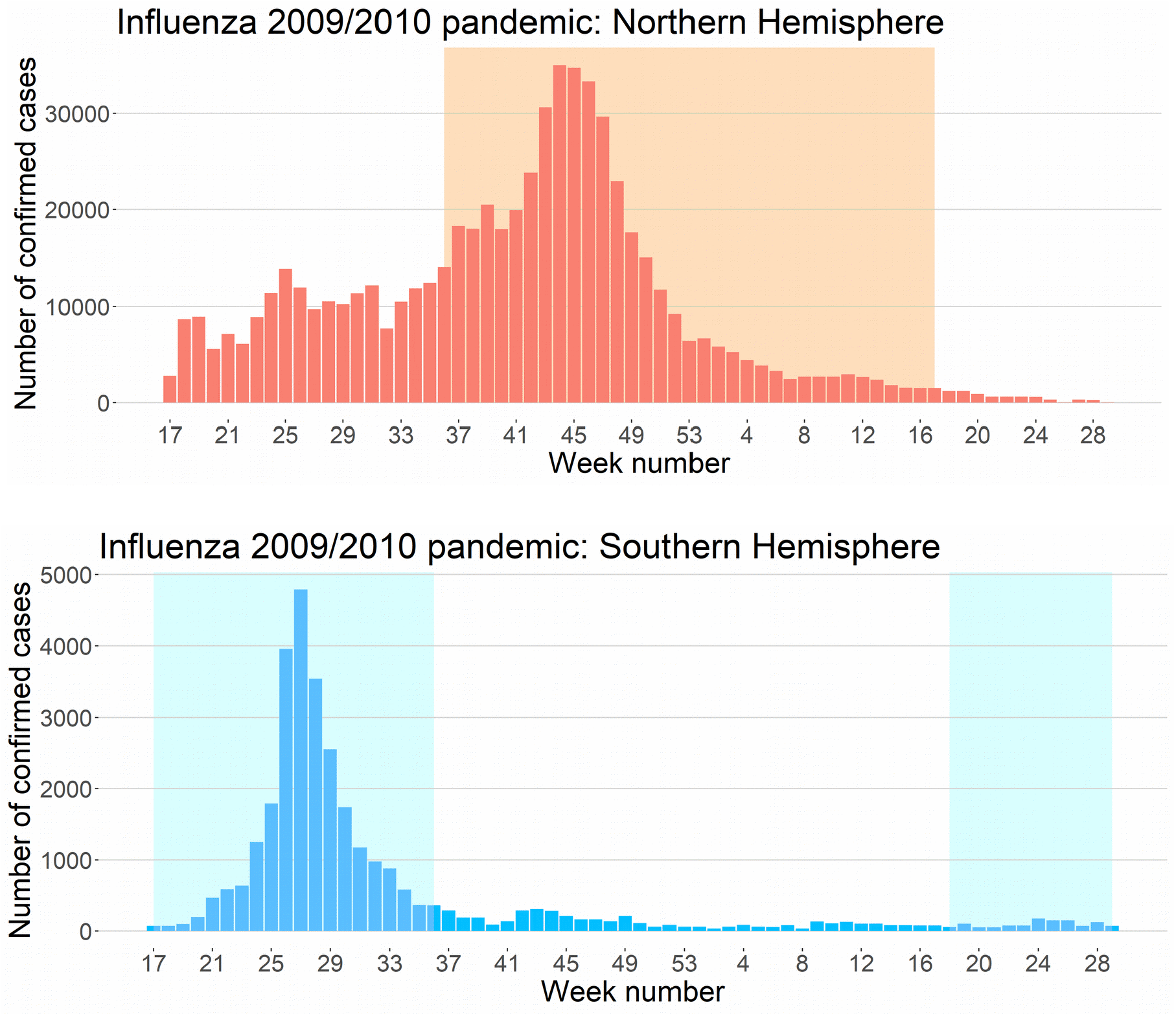
Seasonality in 2009/2010 H1N1 Pandemic: The seasonal pattern can be seen when we compare pooled data from both Hemispheres. High seasonal periods for H1N1 are represented by the shaded areas. In the curve of the Southern Hemisphere, we observe a consistent increase in the number of cases since the beginning of the pandemic, in week 17, which corresponds to the second half of April. In the curve of the Northern Hemisphere, we notice this increase occurs in week 36, which corresponds to the first week of September, although we can already observe an increase in the number of cases in week 41, the first week of October. Likewise, we can expect a second seasonal period for COVID-19 in the Northern Hemisphere starting in September in some countries, with a general spread in October. From the Northern hemisphere curve it can be seen that R0min > 1, but possibly close to 1. Herd immunity has been achieved in most countries and no second wave is seen in the Southern Hemisphere curve after week 17 of 2010.

Seasonality can be directly verified in H1N1 pandemic curves, as we see a different behavior of growth rates along time between the two hemispheres. Influenza pandemic followed a more natural course than the COVID-19 pandemic, where social distancing interventions affected transmission rates, flattening epidemic curves. We can notice that September was the onset of a high seasonal period for the H1N1 pandemic, as there was an increase in the number of cases in the Northern Hemisphere during that month. Similarly, we estimate this period to be the next moment of reversal for the COVID-19 pandemic, and we set April 15 and September 1 as the approximate dates in which the COVID-19 transmission rate will change in the Southern Hemisphere and in the Northern Hemisphere, respectively.

It is important to emphasize that this global seasonal period is an average of the seasonal periods of the countries around the world. It varies from one location to another, and the global seasonal period can be seen as an expected value for a country chosen at random. Therefore, in some places the transmission rate will increase before the expected period, while in others it will increase after the expected seasonal period. For example, in figure 1 we can see that there is a higher increase in the number of cases from the Northern Hemisphere in the beginning of October, which indicates that the seasonal period started later in some countries. Thus, October 1 can be seen as a plausible alternative date for the mean onset of the high seasonal period in the Northern hemisphere.

Note that the asymmetric dates for the onset of high seasons in Northern and Southern hemispheres is reasonable since many factors related with seasonality such as temperature and latitude are not symmetric between both hemispheres.

In addition, it is important to distinguish the seasonal period in which transmission is the greatest from the epidemic period when the number of cases is the highest. Both are highly correlated in seasonal diseases, but the seasonal period usually starts earlier and influences the epidemic period, although the former does not determine the latter. In this sense, many other factors influence the size and duration of epidemic periods, such as the proportion of susceptible populations. This is particularly clear in the Northern hemisphere curve in figure 1, where the number of cases is considerably high from April to August despite being a low seasonal period. However, the increased growth rate after September shows that seasonality increased transmission rate significantly.

The seasonal period begins when epidemics accelerate, that is, we look at the variation in the number of cases and not at the number of cases themselves. Meanwhile, in the epidemic period, we look at the number of cases themselves. Typically, seasonal periods start a few weeks before the epidemic periods and end a few weeks later, especially when the proportion of susceptible population is small and herd immunity was achieved, as we can see in the Southern hemisphere curve of H1N1 2009 pandemic, where mean herd immunity occurred around week 27, 10 weeks before the end of Southern high seasonal period.

### Measuring the Effect of Social Distancing Interventions

We estimate the effect of global social distancing measures because of its intrinsic importance and also to differentiate it from the seasonal effect. This is done because social distancing measures were coincidentally taken at the end of March, which is very close to April, when we expect a change in seasonal periods.

For each of the 50 countries with the greatest epidemics from March 1 to May 1, we collected the dates when the authorities began to adopt social distancing interventions. Further details can be found in the supplementary appendix.

The average initial date for social distancing measures was March 19, with a standard deviation of 6.6 days. Countries in the Northern and Southern Hemispheres had similar initial dates, with both means on March 19. We consider the mean effect of social distancing (MESD) as the difference between the slopes for periods of 10 days before and after the adoption of such measures. Formally, let B be the slope of the regression line from the mean rate of cases from March 17 to March 26, and let A be the slope of the regression line from the mean rate of cases from March 27 to April 5. The one-week gap from March 19 to the beginning of this interval is due to the fact that there is a delay between the adoption of a control measure and its impact on reported cases, since the disease has a median incubation period of four days, and there are some days of delay between the laboratory test and its result. The total delay varies according to the country, but we consider seven days to be a rough estimate of the total delay. The length of the interval is only 10 days because social distancing measures were introduced very close to the expected seasonality period, that should be somewhere in April. Therefore, we must consider it to be as small as possible in order to avoid confusion between the social distancing effect and the seasonality effect. We define MESD = A-B.

Figure 2 shows the graph of the global average of COVID-19 case rates with the regression lines before and after March 19. We obtain slope estimates and 95% confidence intervals given by B= 0.2143 (CI=[0.1720, 0.2567]), and A= 0.0379 (CI= [−0.0135, 0.0893]). MESD= −0.1765, which represents a relative reduction of 82.3%. Note that A and B are not independent. We can suppose that the closer the regression lines are from each other, the more positively correlated A and B are. As the upper limit 0.0893 for A is less than the lower limit 0.1720 for B, we reject the hypothesis that A = B at 95% confidence level. Thus, there is consistent statistical evidence that social distancing measures have decreased, at least in the short term, the global average growth rate of COVID-19 cases with an estimated relative reduction of 82.3% in the speed of growth.

**Figure 2.**
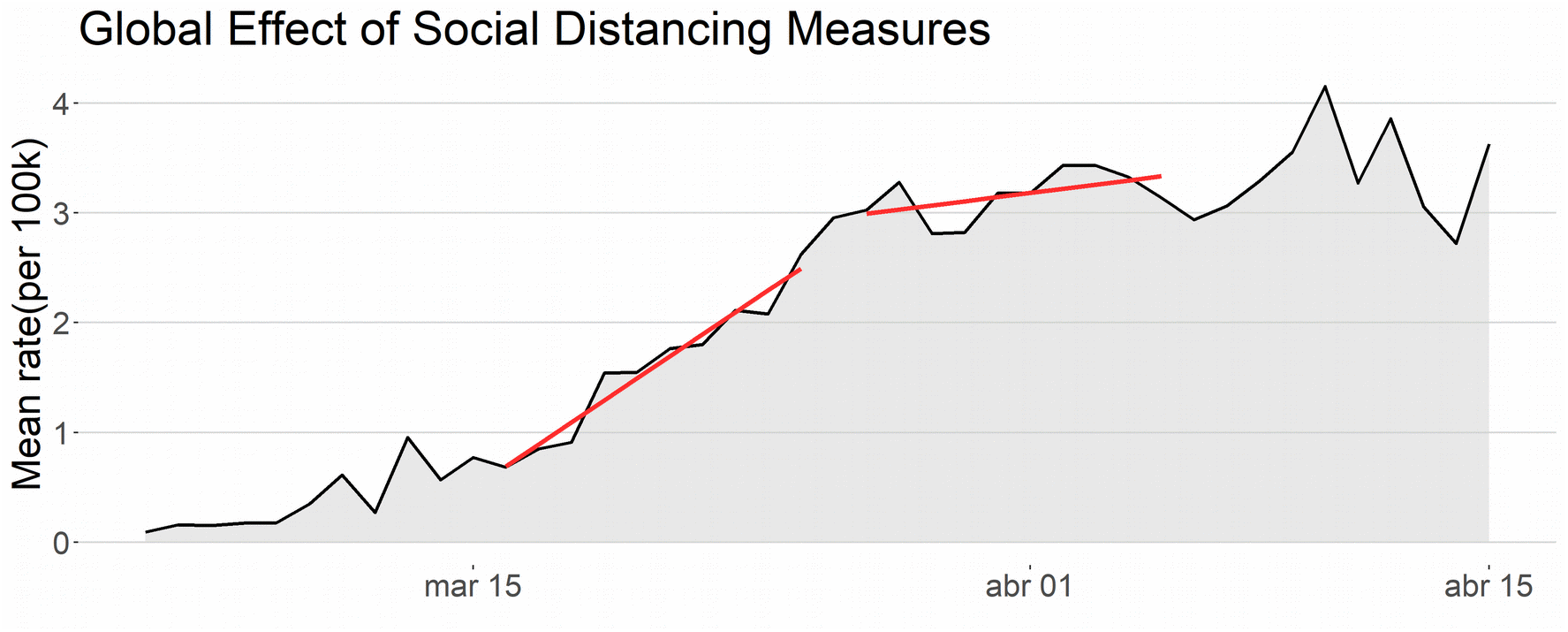
Social distancing effect: The black curve shows the global mean rate of cases per 100k inhabitants. The left red line is the linear regression line for a 10-day period immediately before the effects of social distancing measures were expected to appear. The right red line is the linear regression line for a 10-day period starting one week after March 19, the average initial date for social distancing measures.

In order to ensure that the seasonality effect is not confusing in this analysis, we carried out the same analysis for the Northern and Southern Hemispheres, examining if there is a different behavior in both groups. For the Northern Hemisphere, B=0.2610, A = 0.0431 and MESD= −0.2178, which represents a relative reduction of 83.5%. For the Southern Hemisphere, B=0.0681, A = 0.0130 and MESD= −0.0552, which represents a relative reduction of 81.0%. Hence, the qualitative behavior was the same in both hemispheres during the periods immediately before and after the adoption of social distancing measures, and we conclude that the reduction in the global growth rate at the end of March was not due to seasonality. For further details, see the supplementary appendix.

### Measuring the Seasonality Effect

We are now ready to estimate the effect of seasonality on COVID-19 transmission. First, we consider the seasonal effect for each hemisphere as the variation in the slope for the mean polled rate at the expected moment of seasonal reversal, which we estimate as April 15. As we did to obtain the effect of social distancing, we define the mean seasonal effect in the Northern Hemisphere (MSEN) to be A-B, where B is now the slope of the regression line of the mean rate of cases from March 27 to April 5. Let A be the slope of the regression line of the mean rate of cases from April 16 to May 1. Likewise, we define the mean seasonal effect in the Southern Hemisphere (MSES).

Figure 3 below shows how the mean daily rate of cases changed in different directions, right after the estimated moment of seasonal reversal. To quantify this difference, we obtain the estimates of the slopes of the Northern Hemisphere and 95% confidence intervals given by B=0.0478 (CI=[−0.0138, 0.1094]), A= −0.0586 (CI=[−0.0957, −0.0215]). MSEN= −0.1064, which represents a relative reduction of 222.5%. We interpret a relative reduction greater than 100% as a reduction that changes a positive slope to a negative one. Note that A and B are not independent. We can suppose that the closer the regression lines are from each other, the more positively correlated they are. Since the upper limit −0.0215 for A is less than the lower limit −0.0138 for B, we reject the hypothesis that A = B at 95% confidence level. Thus, there is consistent statistical evidence that the mean seasonal effect in the Northern Hemisphere is smaller than zero (MSES < 0).

**Figure 3.**
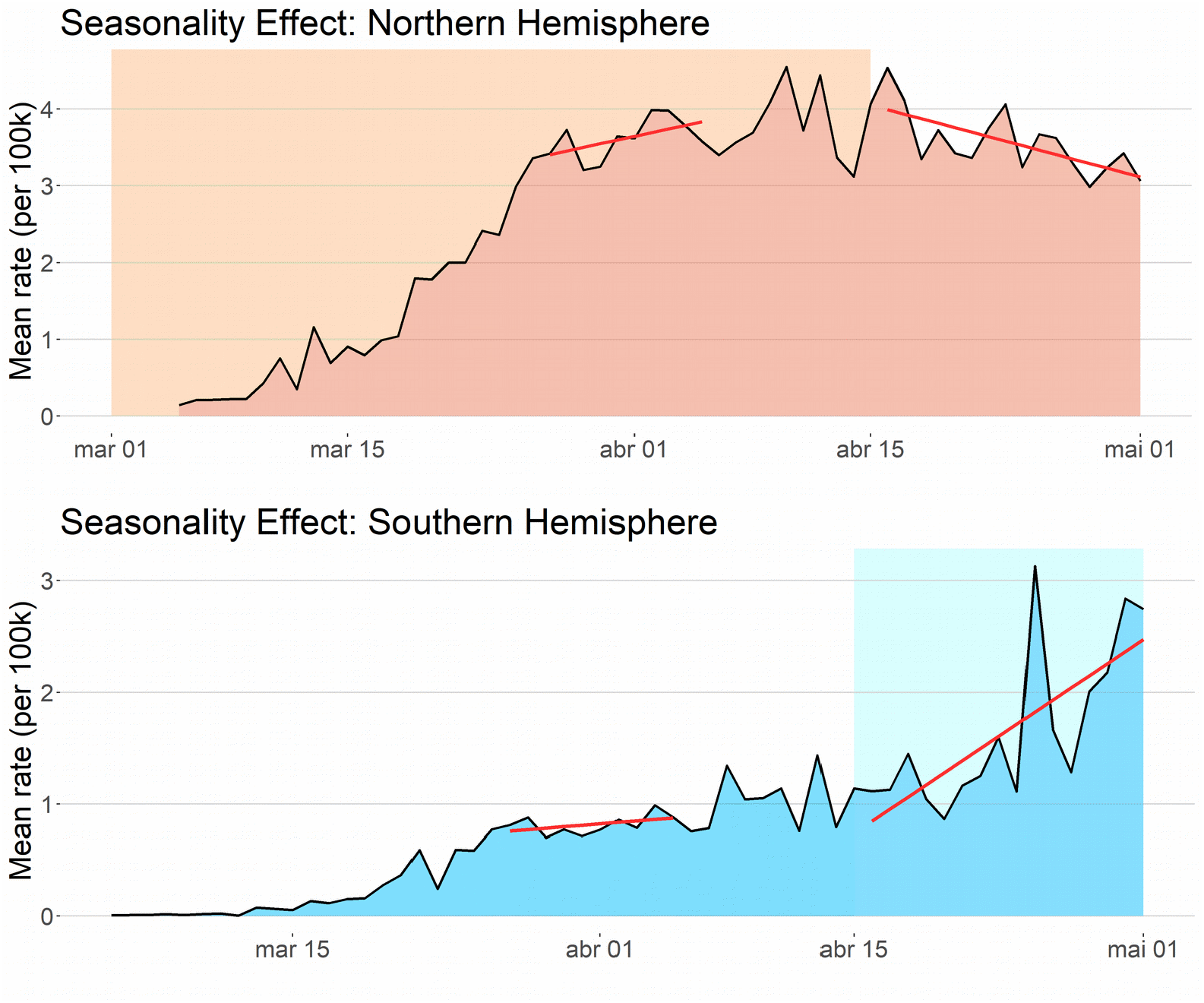
Seasonality effect: black curves show the mean rate of cases per 100k inhabitants for countries in the Northern and Southern Hemispheres, respectively, from March 5 to May 1. The left red lines are the linear regression lines before the expected change of seasonal period. High seasons for Northern and Southern Hemispheres are represented by light-red and light-blue shadow areas, respectively. The right, red lines are linear regression lines immediately after the expected change of seasonal period.

**Figure 4.**
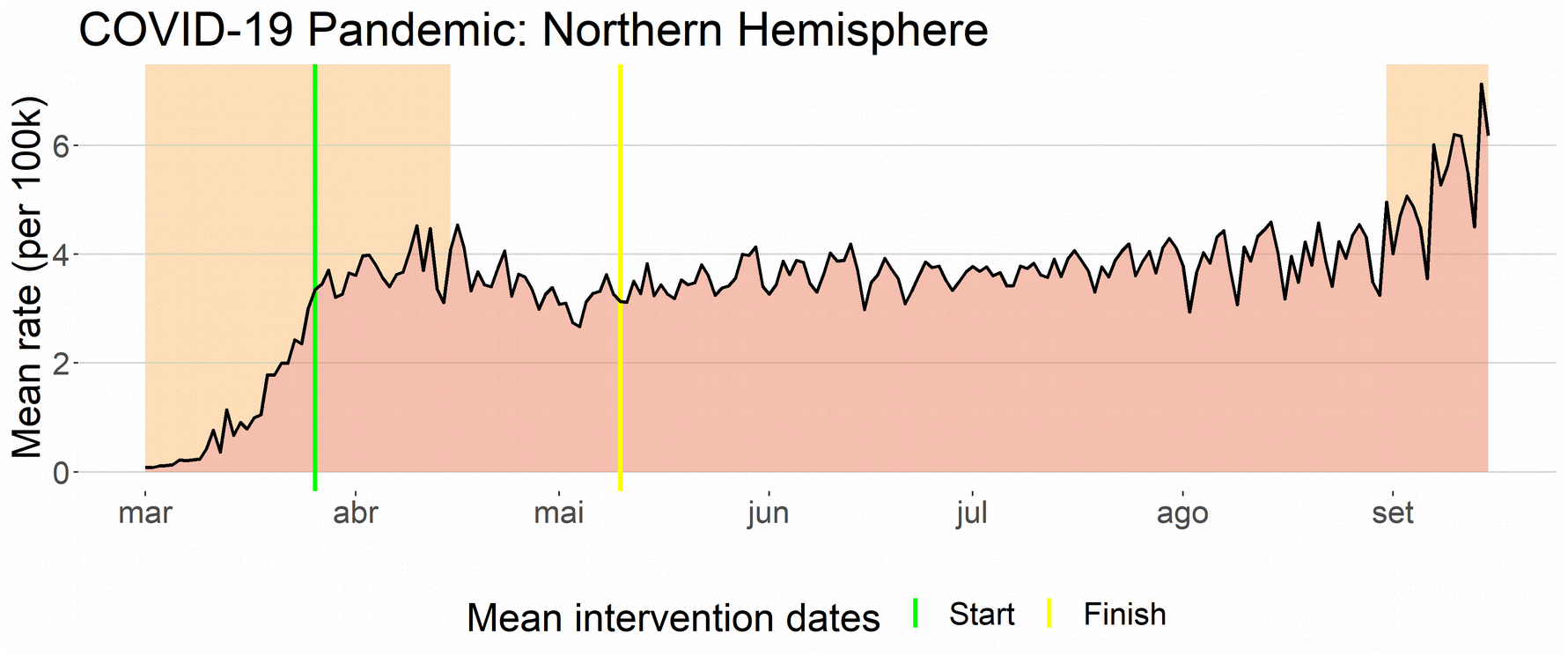
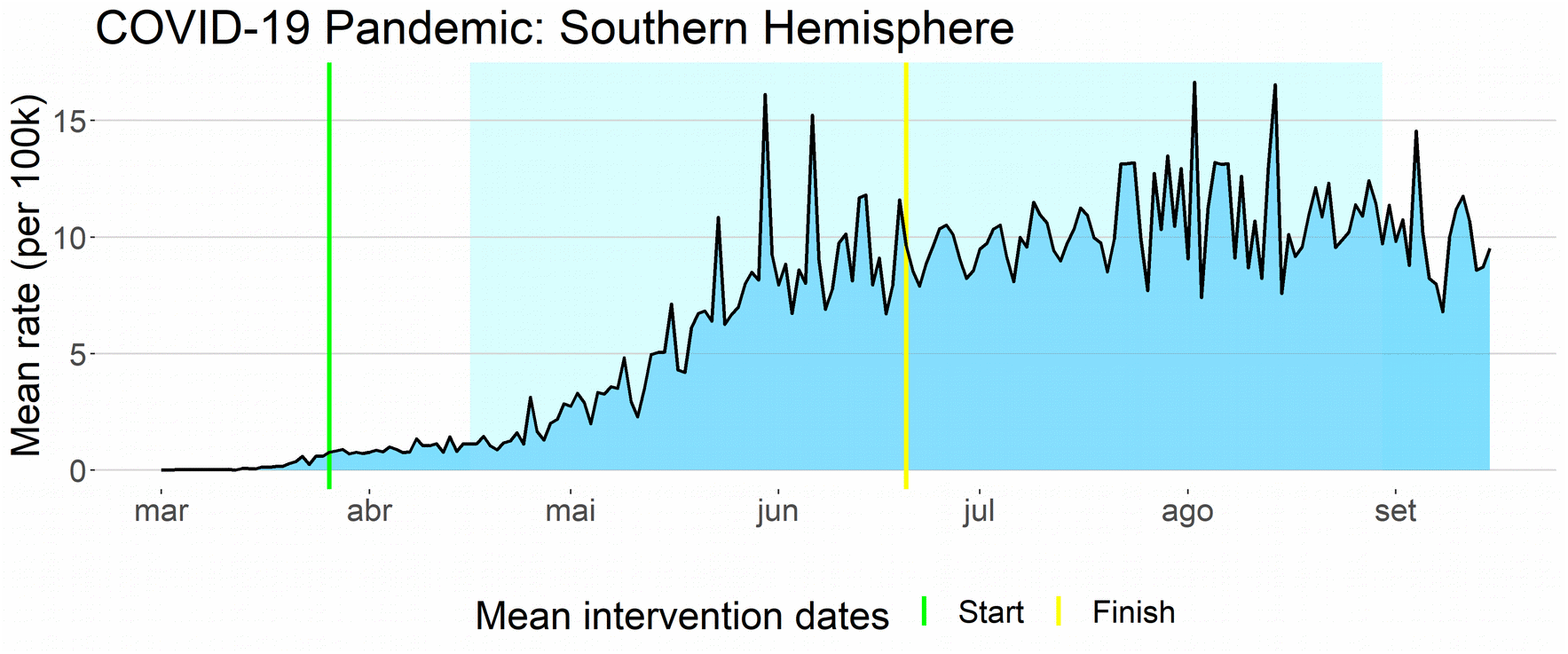
Seasonality pattern in COVID-19 pandemic. Shaded areas represent the high seasonal periods in both hemispheres. Growth rate changes together with the seasonal periods in mid-April and the in September. Between green and yellow vertical lines, social distancing effect decreases the transmission rate. Both social distancing effect and seasonal effect are fundamental to explain the COVID-19 outbreak dynamics.

For the Southern Hemisphere, slope estimates and 95% confidence intervals are given by B=0.0130 (CI=[−0.0081, 0.0341]), A= 0.1089 (CI=[0.0500, 0.1678]). MSES = 0.0959, which represents a relative increase of 740.3%. As the upper limit 0.0341 for A is less than the lower limit 0.0500 for B, we reject the hypothesis that A = B with 95% confidence level. There is consistent statistical evidence that the mean seasonal effect in the Southern Hemisphere is greater than zero (MSES > 0).

To validate the methodology in this database, we verified the sensitivity to time translations of the statistical significance of slope differences in the mean rate curve from March to August. Simultaneous statistical significance for slope differences from both hemispheres is only achieved in mid-April. Details can be found in the supplementary appendix.

Now, consider the plausible hypothesis that the social distancing effect could potentially influence a greater drop in the slope if we consider a period of time greater than the 10 days in its definition. As a consequence, part of the decrease in the slope of the Northern Hemisphere that we attribute to the seasonal effect could be ascribed to the effect of social distancing measures. However, assuming this is true, then a similar decreasing effect would occur in the Southern Hemisphere, and the absolute seasonality effect would be even greater. This is a contradiction since we would have a very small seasonal effect in one hemisphere and a very large one in the other.

In addition, consider the hypothesis that a fatigue effect could explain the increase in the mean rate of the Southern Hemisphere. The average time interval between the beginning of social distancing measures (March 19) and the beginning of the data time interval used to measure the seasonal effect (April 16) is less than a month. Therefore, it seems unlikely that the effect of fatigue could be responsible for the increase in the rate of cases in the Southern Hemisphere and in such a short time. Besides, we would again have an opposite effect in the Northern Hemisphere, which seems to be a contradiction. Another possible confounding factor, the social-economic one, is analyzed in detail in the supplementary material. These factors and others that could possibly affect rates in reported data, such as heterogeneous transmission, spatial spread in meta-population models and increased rate of testing, typically produce similar effects across countries, mostly in large groups such as the Northern and Southern Hemispheres. Although quantitative differences are expected for these factors, a similar qualitative effect is likely to occur in both hemispheres.

In summary, slope differences between close time intervals like the one we used to define seasonal effect may occur for a number of reasons in epidemiological data. What differs the seasonal effect from all those reasons appointed above is that seasonality produces synchronous opposite effects in the Northern and Southern Hemispheres. Furthermore, for COVID-19 pooled data this effect was considerably large in both hemispheres and occurred precisely in the estimated onset of seasonal period of the Southern Hemisphere. Hence, there is sufficient statistical evidence that shows a consistent seasonality effect in the COVID-19 pandemic.

In order to confirm that the difference between the growth speeds from the end of April and the end of March is due to seasonality and not to confounding effects, we run a multiple linear regression analysis. We take as response variable Y the seasonality effect (A-B) of each country. The explanatory variables are the seasonal factor X_HP_, the social distancing factor X_SD_ and the income factor X_IC_. We set X_HP_ as the indicator variable if the country belongs to the Northern Hemisphere, X_SD_ as the discrete score varying from 0 to 2 according to the level of social distancing measures adopted (low/none, moderate, high/lockdown) and X_IC_ as the country’s gross domestic product per capita (GDP). After rescaling variables to a comparable scale, we obtained for the additive model the estimate Y = 0.20 −0.18X_HP_ −0.06X_SD_ −0.09X_IC_. All of the three factors contribute to a reduction on Y, but the seasonal factor X_HP_ had the largest absolute effect.

To access dependence between factors, we run a multiple linear regression model with interaction terms, obtaining for re-scaled variables the estimate Y = 0.11 −0.16X_HP_ +0.03X_SD_ +0.01X_IC_ −0.01X_HP_X_SD_ −0.01X_HP_X_IC_ − 0.09X_SD_X_IC_. Note that the absolute value of the individual coefficient of the seasonal factors X_HP_ remains the largest, and its estimate is very close to the estimate obtained in the additive model. The interaction coefficients between the seasonal factor and the two other factors are considerably small, which shows that the seasonal factor affected all the countries, regardless of the levels of social distancing and income factors. The individual coefficients for the social distancing and income factors changed from negative to positive, which shows that these factors did not contribute individually to reduce the growth rate of cases. The reduction measured previously in the additive model is contained in the interaction between the social distancing factor and the income factor. This implies that social distancing effectiveness is highly correlated with income and its impact was bigger in high-income countries than in low-income ones. Details and further regression analysis can be found in the supplementary appendix.

### Modeling COVID-19 Pandemic

COVID-19 pandemic was unique in its dynamics, mostly because of the worldwide adoption of social distancing interventions. We propose a seasonal SEIR model with an additional parameter δ that represents the social distancing effect. We take δ as a constant number that decreases β(t) during the time interval in which the interventions are adopted. In this model, seasonal effect and social distancing effect drive the dynamics of the outbreak. Details can be found in section 5.3 of the supplementary appendix.

We display below, the mean rates for both Hemispheres from March 1 to September 15. Seasonal periods are represented by shaded areas and mean dates of social distancing interventions are represented by vertical lines. As in our simulation results, we can see that significant changes in transmission rate for COVID-19 pandemic is driven by seasonal forces and social distancing effects.

## 3. Discussion

In this study, we demonstrate that seasonality strongly affects COVID-19 transmission and that its seasonal period follows the autumn/winter pattern that is typical of other viruses with respiratory transmission. We also measured the social distancing effect, providing the distinction between the seasonal effect, the social distancing effect and the income effect.

Our results partially contradict the results obtained by Flaxman et al.^12^ and Islam et al.^13.^ In the first study, the decrease in daily cases in 11 European countries in late April and early May is attributed exclusively to social distancing interventions without considering seasonality or other possible confounding factors. In the second one, worldwide data are analyzed, but again the entire effect is attributed solely to social distancing interventions without considering other factors such as seasonality.

### Prediction of Second Waves and Consequences for Decision-making

The Seasonal force of transmission drives the second waves in seasonal diseases. Once the seasonal effect is established in COVID-19 and its seasonal periods are known, we can predict the appearance of future outbreaks. In particular, further epidemic waves are expected to occur from September to February in the Northern Hemisphere. In contrast, a significant reduction in the transmission rate is expected to occur in countries of the Southern Hemisphere in the same period.

High seasonal periods are time intervals when the transmission rate is the highest in each hemisphere. Nevertheless, we must remember that epidemic periods also depend on other variables, such as the percentage of susceptible population, and that each city has its own seasonal transmission rates β_max_ and β_min_. Thus, places that have been affected by major epidemics will be less impacted by this change in the transmission rate. Possible examples include Sweden and some cities in the United States that have already experienced major epidemics and where the increase in the transmission rate will have less impact on the size of the epidemic due to a smaller proportion of susceptible population. On the other hand, some countries with very large proportions of susceptible population will be more affected if they do not control their epidemics. Some examples include most countries in Europe.

The first wave in most of Europe and the rest of the Northern Hemisphere generally lasted only two months or less, from mid-February to mid-April. Social distancing measures had to be adopted for around two months before the seasonal period ended. In May, when most European countries started to ease their social distancing measures, the transmission rate was lower and the epidemics were controlled with less stringent measures. Cities with R0_min_ < 1 did not witness the continuation of the epidemic. Other locations, such as some cities in the United States, certainly have R0_min_ > 1 and, therefore, the epidemic continued to increase. Nonetheless, it is important to note that they have also been affected by the seasonal effect.

In the Southern Hemisphere, social distancing measures had an effect similar to that in the Northern Hemisphere, producing a stabilization in the number of cases in the first two weeks of April. However, the seasonal effect produced a significant increase in the transmission rate, which went from β_min_ to β_max_, and with the implementation of social distancing measures, the number of cases increased consistently from April 15. Also, more restrictive measures such as lockdowns, though effective, could not be adopted for the entire seasonal period that lasts over six months. An example is Argentina that kept its epidemic controlled by lockdown measures similar to those in Europe. However, when it started to lift its restrictions, either officially or due to population fatigue, the number of cases increased, unlike what happened in Europe, because the transmission rate was β_max_ instead of β_min_. From October 2020 to March 2021, the transmission rate in the Southern Hemisphere will decrease from β_max_ to β_min_. We expect the epidemic to be controlled in places where Rt < 1. Nevertheless, some countries with large proportions of susceptible populations, such as New Zealand and Australia, may experience major epidemics in cities where Rt > 1, if no control measures are adopted.

Taking into account the seasonality effect, we could have predicted since May 2020 that many, though not all of the countries in the Northern Hemisphere, will experience second waves that will generally begin in September and increase in October and November. In fact, in September 2020, when this article was finalized, there was an increasing number of cases in some countries in the Northern Hemisphere. Restrictive social distancing measures such as lockdowns may not be as effective as they were in the first wave because the high-season period will last an entire six-month span. Other control measures, such as vaccinating part of the population, must be implemented. Until vaccines are available, other control measures, such as active contact tracing, will have to be adopted; otherwise, effective social distancing will have to last until either a vaccine is available or mid-April. Other alternatives are mitigating measures, such as less restrictive social distancing and waiting for herd immunity.

Seasonality plays a fundamental role in vaccine scheduling.^14^ Therefore, public health officials should take this in consideration when a vaccine is available for COVID-19. The immunization of the world population will be more effective if applied between February and April in countries of the Southern Hemisphere and between August and October in countries of the Northern hemisphere, although we emphasize that these dates are averages and public health officials must take into account the specific seasonal periods of their specific country.

### Detection of Seasonality in Future Pandemics

We briefly describe below the steps needed to detect the presence of seasonal effects in future pandemics:

1. Comparing aggregated slopes in the Southern and Northern Hemisphere curves can be used to detect seasonality at the onset of a pandemic, even with data from a short period of time. In this preliminary analysis, attention to confounding factors must be taken into consideration.
2. Monitoring abrupt changes in their slopes (either R0_t_ or β_t_) close to the onset of a high seasonal period is a way to confirm this detection.
3. Measuring changes in data of the chosen parameters before and after the onset of a high seasonal period provides an estimate of the seasonal effect.

Our method is particularly useful to detect seasonality during a pandemic, since a large amount of data from both hemispheres is available in this case. In minor epidemics, data from many countries are not available, but those from a small number of countries can still be used to apply this methodology as long as data from countries in both hemispheres are available.

## Data Availability

All data used in this manuscript is public available.

## Acknowledgments

The author would like to thank Rodrigo M.C. Dias for his support with the data and figures, Cintia Shimokomaki, Kyoko Shimokomaki and Laura L. Alves de Souza for a full reading of the mnuscript and Fernanda Di Genio for fruitful discussions.

## Data and materials availability

The main data was extracted from the John Hopkins University website https://coronavirus.jhu.edu/. More details and information are available in the supplementary appendix.

