## Supplementary Appendix for "Early detection of seasonality and second-wave prediction in the COVID-19 pandemic"

Márcio Watanabe

**Table of contents**

1. Methods…....…..…………………………………………..………………………………….…..…………....2

2. Social distancing effect…………………………………......………………………………………..…..…....4

3. Total number of confirmed cases……………….……………..…..………………………………………......8

4. Confirmatory analysis of seasonal effect

4.1 Methodology validation.….……..…………………………………………………………………..10

4.2 Seasonal mean rate time series………………………………………………...………………..12

4.3 Subgroup analysis: low-income countries……...…..…………………………...……………...…13

4.4 Multiple linear regression analysis ……….…………………………...……………………………..14

5. Modeling the seasonality effect

5.1 Seasonal SEIR model……..……………………………………...…………………………..… 16

5.2 Seasonal pattern in COVID-19 data..……………………………………………………………...19

5.3 Modeling COVID-19 outbreaks……..……..………………………………………………………...21

6. Limitations

6.1 Reported cases in China…….……………………………………….……..……………….…..26

6.2 Regional seasonal patterns.….……...…..…………………………………………………………….27

6.3 Case definitions……………………………………………………………...………….……....28

Supplementary references…………....…………………………..…………………………………….……....30

#### 1. Methods

This section is a summary of the methodology applied in this study. Further details can be found in the ensuing sections. The data we use were obtained from the John Hopkins University website. The data were analyzed using the statistical software R 64 bits version 4.0.0

We gathered daily data from the 50 countries with the greatest epidemics, as measured by the number of confirmed cases from March 1, 2020 to May 1, 2020. The inclusion criteria were: 1) having at least 1000 confirmed cases in this period; 2) having data of daily confirmed cases either by notification or by the onset of symptoms; 3) having at least 90% of its territory in the same hemisphere. This last criterion was necessary in order to clarify the hemisphere to which a country would be assigned. Based on this, Colombia, Ecuador, Indonesia and Singapore were removed from the pool. The advantage of restricting the number of countries in the pool, instead of using the largest possible number, is to avoid the inclusion of very small outbreaks, which are more susceptible to random variation, and also to exclude countries without an established national health surveillance system.

Then, whenever appropriate, we separate countries into two different groups, according to whether most of their territory is in the Northern or Southern Hemispheres. We obtained insufficient countries for the Southern Hemisphere group, so we decided to include other countries that had more than 1000 cases in the period analyzed and that were among the top 100 epidemics. Using these criteria, Argentina and New Zealand were included in the Southern Hemisphere group. For each country, we calculate the rate of confirmed cases per 100k inhabitants. For each day, we calculate the rate per 100k inhabitants for each selected country and also its simple mean. We study the global dynamics of the pandemic by analyzing the time-series curves for mean daily cases and rates.

We can interpret the time-series curves of the mean rates as showing the mean dynamics of the epidemic in each hemisphere from a country perspective. That is, each country has the same weight to the mean curve. An advantage of this curve is that it gives us an insight into the behavior of the pandemic itself, without being overly affected by a single country. Time series of daily number of cases or daily rates are more suited to compare seasonal effects than time series of cumulative number of cases, since the latter produces monotonically increasing curves and the information of growth variation is more easily seen in non-monotone curves.

Seasonal effects can be measured in a variety of ways. We believe there are many advantages in pooling data from different countries and dividing these into the Northern and Southern Hemisphere groups to measure the seasonal effect. One reason is that we consider seasonality as a single complex phenomenon that is influenced by many factors, instead of reducing it to a single factor, such as temperature. In addition, integrating data increases the statistical power of the analysis.

We define the expected moment of seasonality reversion as the expected week, day or month when the transmission rate of a seasonal disease changes due to seasonal reasons. This variable varies from country to country, and even within a country itself. It can also vary from one disease to another. Since COVID-19 is transmitted as a respiratory disease, we estimate the expected seasonality moment of reversal for COVID-19 based on the 2009 H1N1 pandemic and data from other respiratory syndromes.

To quantify the change in confirmed case rates in each hemisphere, we perform a simple linear regression before and after the expected moment of seasonality reversal. To decrease the influence of other factors and a possible confounding effect, we consider the slopes for a short period of ten days before and two weeks after the expected seasonality moment of reversal.

The mean seasonal effect for the Northern Hemisphere (MSEN) is calculated as the difference between the slopes for this hemisphere before and after the expected seasonality moment of reversal. We test the hypothesis that seasonality did not affect COVID-19 transmission with the null hypothesis H0: MSEN=0. In the same manner, we perform definition and testing for the Southern Hemisphere.

We calculate the slope variation for each hemisphere at time point t as the difference between the slope of the curve in a short period of days (ten days or two weeks) after t minus the slope of the curve in a short period of days before t. The 10-day (2-week) extension was chosen because a shorter time interval would be subject to a great variability due to a small number of time points in its estimation. On the other hand, a longer period of time would be subject to the influence of other factors, generating possible confounding factors.

Slope differences for different time intervals naturally occur in epidemic curves. For example, herd immunity is expected to decrease growth speed as the epidemic evolves. To assess the statistical significance of the methodology, we display the daily difference between the bounds of confidence intervals for the two slopes and verify other dates when this slope difference was also significant at 95% confidence level.

Epidemic curves of single outbreaks are expected to have initially approximate exponential growth. However, we are pooling data from a large number of countries, so in a meta-population like the one in this study an exponential approximation is no longer valid (1). Since we are measuring the seasonal effect in a short period of time, the linear approximation produces reasonable results.

To corroborate that the seasonality effect is due to seasonality and not to confounding factors, we run a multiple linear regression analysis. We take as response variable Y the seasonality effect (A-B) for each country. The explanatory variables are the seasonal factor $X_{HP}$, the social distancing factor $X_{SD}$ and the income factor $X_{IC}$. We set $X_{HP}$ as the indicator variable if the country belongs to the Northern Hemisphere, $X_{SD}$ as the discrete score varying from 0 to 2 according to the level of social distancing measures adopted (low, moderate, high/ lockdown) and $X_{IC}$ as the country's gross domestic product per capita (GDP).

To assess the variability of the seasonal effect between different countries, we display separately the boxplots of the seasonal effects (slope differences before and after the expected seasonality moment of reversal) for countries from both hemispheres.

A modified seasonal SEIR compartmental model is introduced to model COVID-19 outbreaks. The modification includes a multiplying factor to account for the social distancing effect. Simulation results are compared to data from COVID-19 outbreaks.

Main limitations of the methodology and database are discussed.

#### 2. Social distancing effect

In this section, we display more details about the analysis of the effects of the social distancing measures.

In Table 1 below, we display the top 50 countries with the largest number of confirmed cases from 2020-03-01 to 2020-05-01, in addition Argentina and New Zealand. We also show the social distancing intervention dates in each country. For countries that adopted different social distancing measures on different dates, we considered only the first one. Belarus, Japan, South Korea and Sweden did not widely adopt social distancing interventions in this time period and thus were excluded from this specific analysis.

**Table 1. Dates of Social Distancing Interventions**

| Country | First initial date | Reopening date |
|---|---|---|
| Argentina | 2020-03-20 | - |
| Australia | 2020-03-23 | 2020-07-01 |
| Austria | 2020-03-16 | 2020-05-01 |
| Bangladesh | 2020-03-26 | 2020-06-01 |
| Belarus | - | - |
| Belgium | 2020-03-18 | 2020-05-04 |
| Brazil | 2020-03-24 | 2020-06-16 |
| Canada | 2020-03-23 | 2020-05-19 |
| Chile | 2020-03-19 | 2020-07-19 |
| Colombia | 2020-03-25 | 2020-09-01 |
| Czech Republic | 2020-03-16 | 2020-04-27 |
| Denmark | 2020-03-13 | 2020-04-15 |
| Dominican Republic | 2020-03-19 | 2020-07-01 |
| Ecuador | 2020-03-16 | 2020-05-11 |
| Egypt | 2020-03-25 | 2020-04-24 |
| Finland | 2020-03-16 | 2020-04-15 |
| France | 2020-03-17 | 2020-05-11 |
| Germany | 2020-03-23 | 2020-05-06 |
| India | 2020-03-25 | 2020-05-04 |
| Indonesia | 2020-03-28 | 2020-06-01 |
| Iran | 2020-03-14 | 2020-05-03 |
| Ireland | 2020-03-12 | 2020-05-18 |
| Israel | 2020-04-02 | 2020-04-19 |
| Italy | 2020-03-09 | 2020-05-04 |
| Japan | - | - |
| Malaysia | 2020-03-18 | 2020-05-04 |
| Mexico | 2020-03-23 | 2020-05-18 |
| Netherlands | 2020-03-15 | 2020-05-11 |
| New Zealand | 2020-03-10 | 2020-05-14 |
| Norway | 2020-03-12 | 2020-04-27 |
| Pakistan | 2020-03-24 | 2020-05-09 |
| Panama | 2020-03-25 | 2020-05-13 |
| Peru | 2020-03-16 | 2020-07-01 |
| Philippines | 2020-03-15 | 2020-06-01 |
| Poland | 2020-03-13 | 2020-04-19 |

| | | |
|---|---|---|
| Portugal | 2020-03-19 | 2020-05-04 |
| Qatar | 2020-03-11 | 2020-06-15 |
| Romania | 2020-03-25 | 2020-05-15 |
| Russia | 2020-03-28 | 2020-05-12 |
| Saudi Arabia | 2020-03-09 | 2020-05-28 |
| Serbia | 2020-03-15 | 2020-05-05 |
| Singapore | 2020-04-07 | 2020-05-05 |
| South Africa | 2020-03-26 | 2020-05-01 |
| South Korea | - | - |
| Spain | 2020-03-14 | 2020-05-11 |
| Sweden | - | - |
| Switzerland | 2020-03-16 | 2020-04-27 |
| Turkey | 2020-04-03 | 2020-05-11 |
| Ukraine | 2020-03-17 | 2020-05-12 |
| United Arab Emirates | 2020-03-26 | 2020-04-24 |
| United Kingdom | 2020-03-23 | 2020-05-13 |
| United States | 2020-03-19 | 2020-06-12 |
| Northern Hemisphere mean | 2020-03-19 | 2020-05-10 |
| Southern Hemisphere mean | 2020-03-19 | 2020-06-13 |
| Global mean | 2020-03-19 | 2020-05-17 |

The Global mean included all of the countries from the table, except Belarus, Japan, South Korea and Sweden. The Southern Hemisphere mean represents the average of the intervention dates from Argentina, Australia, Brazil, Chile, Peru, South Africa and New Zealand. The Northern Hemisphere mean was obtained from the Global group excluding the following countries from the Southern Hemisphere group, Colombia, Ecuador, Indonesia and Singapore, as they were not assigned neither to the Northern Hemisphere group nor to the Southern Hemisphere one because they did not fit into one of the inclusion criteria (i.e., their territories are mostly close to the Equator line).

Both the Northern and Southern Hemisphere groups have the same truncated initial intervention mean date 2020-03-19 (Southern: mean = 19.7; standard deviation = 5.4), (Northern: mean = 19.5; standard deviation = 6.3). The Global truncated initial intervention mean date, which also includes the four countries close to the Equator line, is 2020-03-19 (mean = 19.8; standard deviation = 6.6). The initial (first) interventions dates were considerably synchronized across the world.

However, the reopening dates of social distancing interventions vary significantly worldwide. Besides, it is difficult to establish a single criteria to define one reopening date since each country relaxed its social distancing rules in different phases. In addition, large countries adopted different dates for different regions. We have tried to choose, whenever possible, the date were the relaxation could possible impact the transmission the most. For example, in the United States and Australia the mean date of reopening bars was the relaxation which impacted transmission the most. The mean and standard deviation of the chosen dates as the reopening were: World group (mean = 2020-05-17; standard deviation = 26.7); Northern Hemisphere (mean = 2020-05-10; standard deviation = 16.7); Southern Hemisphere (mean = 2020-06-13; standard deviation = 30.6).

In the main text, we have shown results for the global mean daily rates (see Figure 1). Next, in Figure S1 below, we show the mean rates for the Northern and Southern Hemisphere groups from 2020-03-05 to 2020-04-15. It also shows the regression lines before and immediately after the expected effect of social distancing interventions. We estimate an approximate 7-day delay from the mean intervention date 2020-03-19 and a possible effect on the pooled rate due to the time of symptom onset as well as testing results. Thus, the left regression lines (before intervention) use data from March 17 to March 26, while the right regression lines use data from March 27 to April 5.

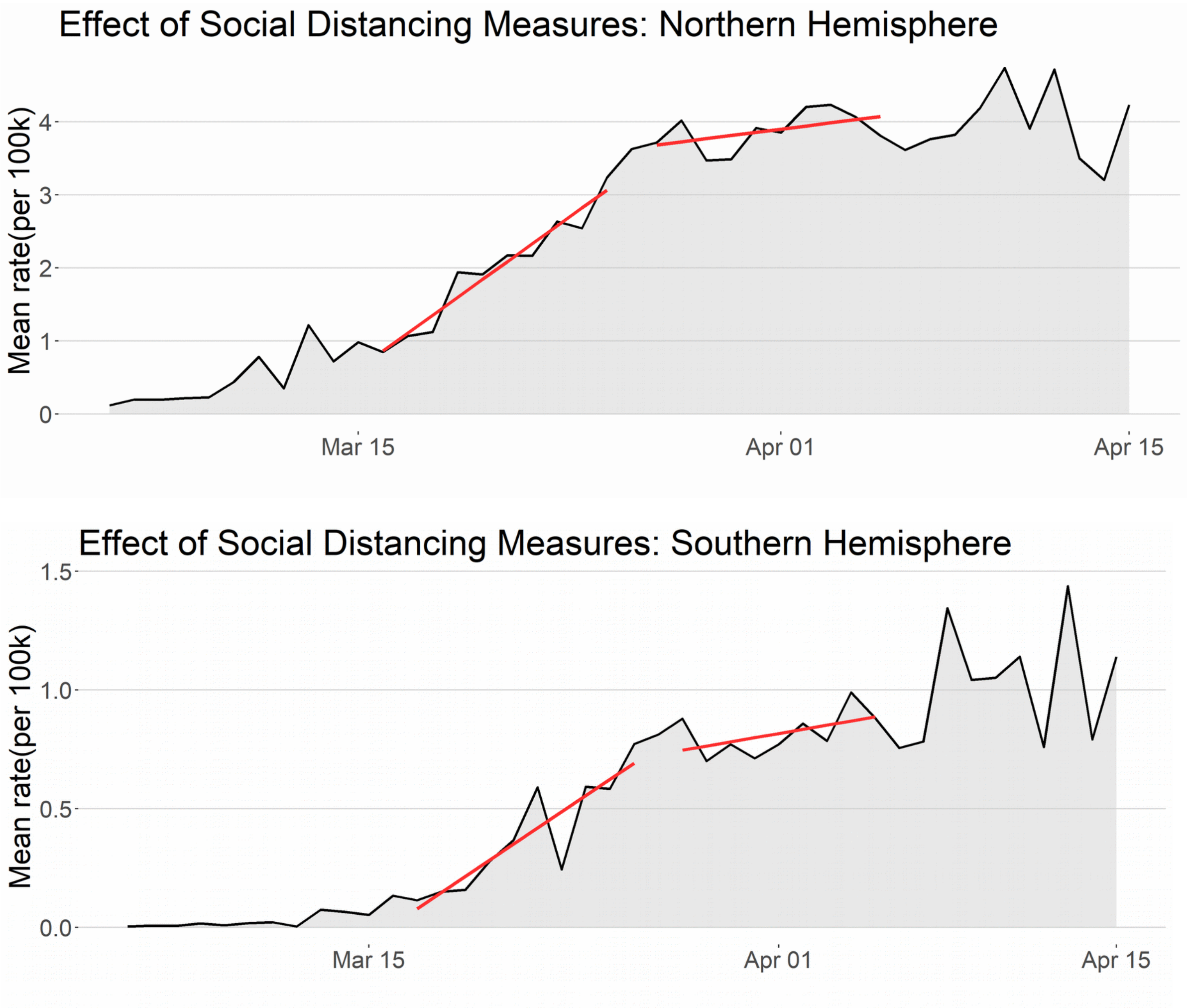


**Figure S1 - Social distancing effects:** The black curves show the mean rates of cases per 100k inhabitants in the Northern and Southern Hemispheres, respectively. The left, red lines are the linear regression lines for a period of 10 days before the effects of social distancing measures appear. The right, red lines are the linear regression lines for a 10-day period starting one week after March 19, the average initial date for social distancing measures.

We observe similar qualitative behavior in both hemispheres. The curves of both hemispheres had a decrease in their growth speeds measured by the difference between slopes from the right and left regression lines. To measure the quantitative effect, we obtain slope estimates for both regression lines before (B) and after (A) and 95% confidence intervals.

For the Northern Hemisphere, we have B=0.2610 (CI= [0.2048, 0.3172]), A=0.0431 (CI= [-0.0193, 0.1055]). MESD= -0.2178, which represents a relative reduction of 83.5%. Note that A and B are not independent. We can suppose that the closer the regression lines are from each other, the more positively correlated A and B are. As the upper limit 0.1055 for A is less than the lower limit 0.2048 for B, we reject the hypothesis that A = B at 95% confidence level. Thus, there is sufficient statistical evidence that the social distancing measures have decreased, at least for the short term, the Northern average growth rate of COVID-19 cases with an estimated relative reduction of 83.5% in the speed of growth.

For the Southern Hemisphere, we have B=0.0681 (CI= [0.0393, 0.0969]), A=0.0130 (CI= [-0.0081, 0.0341]). MESD= -0.0552, which represents a relative reduction of 81.0%. We suppose that A and B are positively correlated. As the upper limit 0.0341 for A is less than the lower limit 0.0393 for B, we reject the hypothesis that A = B at 95% confidence level. Thus, as previously stated, there is sufficient statistical evidence that the social distancing measures have decreased, at least for the short term, the Southern average growth rate of COVID-19 cases with an estimated relative reduction of 81.0% in the speed of growth. Note that the relative reductions in both hemispheres are very similar.

#### 3. Total number of confirmed cases

In the main text, data analysis was based on rates (number of cases per 100k inhabitants). Rates provide a country's perspective of the distribution of the epidemics, in the sense that each country has the same weight in the mean calculation. We repeat here the analysis of the seasonality effect for the total number of cases and not for the mean of rates. This provides a population perspective to the analysis, in the sense that each person has the same weight in calculations. This can be interpreted as having only two countries in the world, the Northern and Southern Hemispheres, and we were comparing their epidemics. One of the disadvantages of the next analysis is that outliers have a stronger influence here than in the mean rate analysis.

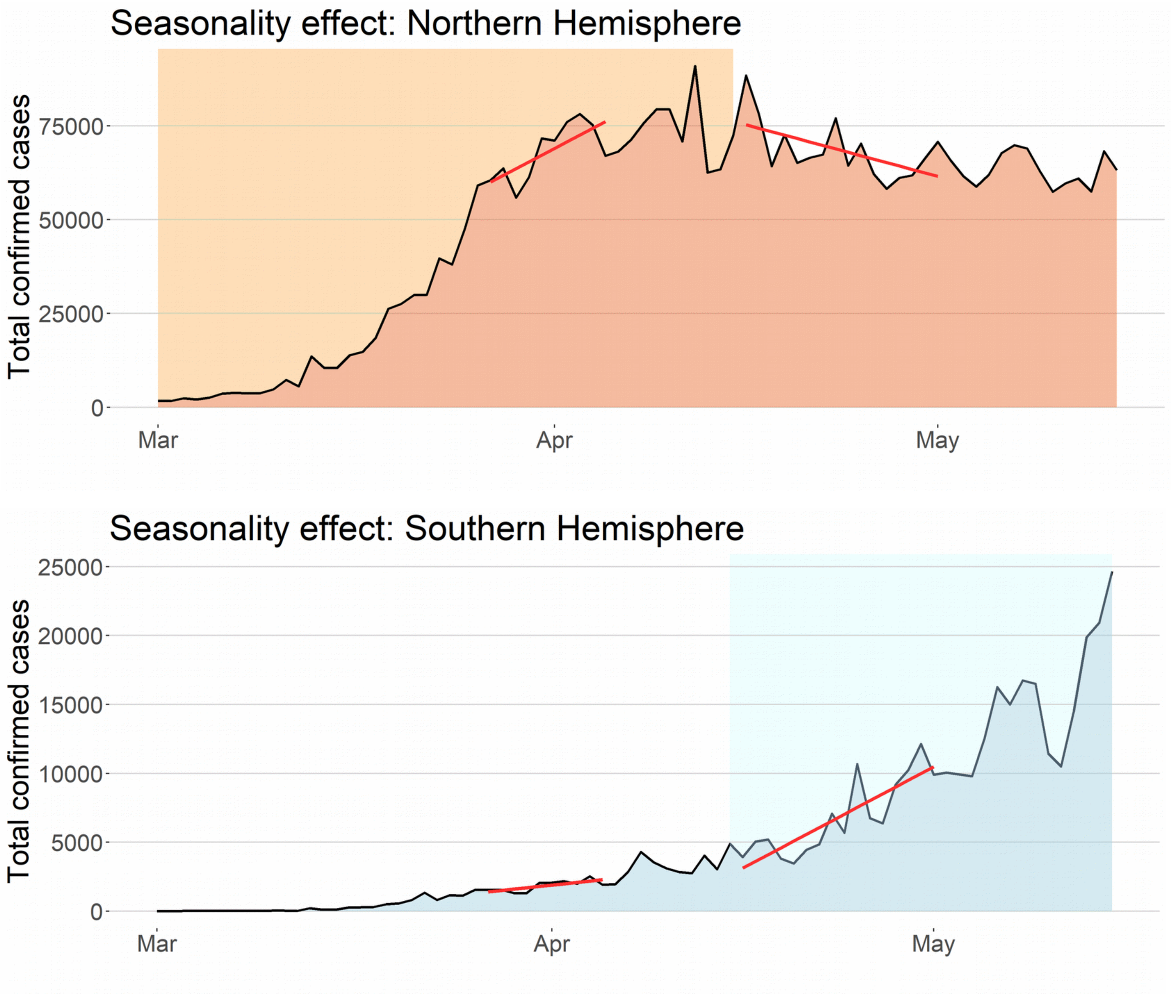


**Figure S2 - Seasonality effect:** High seasons are represented by shaded areas. The black curves show the total number of confirmed cases in the Northern and Southern Hemispheres, respectively. The left, red lines are the linear regression lines immediately before the expected seasonal reversal moment, and the right, red lines are the linear regression lines immediately after the expected seasonal reversal moment for both hemispheres.

Figure S2 above gives a clear picture of how the total number of cases moved into different directions in the Northern and Southern Hemispheres immediately after the estimated moment of change in the seasonal period. It has the same qualitative behavior of the mean rates for both hemispheres.

To quantify this difference, we obtain the estimates of the slopes of the Northern Hemisphere and 95% confidence intervals given by B=1787.7 (CI=[374.5, 3201.0]), A= -914.1 (CI=[-1677.6, -150.6]). MSEN= -2701.8, which represents a relative reduction of 151.1%. We interpret a relative reduction greater than 100% as a reduction, which changes a positive slope to a negative one. Note that A and B are not independent. We can suppose that the closer the regression lines are from each other, the more positively correlated A and B are. As the upper limit -150.6 for A is less than the lower limit 374.5 for B, we reject the hypothesis that A = B at 95% confidence level.

For the Southern Hemisphere, slope estimates and 95% confidence intervals are given by B=97.3 (CI=[24.3, 170.2), A= 490.5 (CI=[310.6, 670.4]). MSES= 393.2, which represents a relative increase of 404.1%. As the upper limit 170.2 for B is less than the lower limit 310.6 for A, we reject the hypothesis that A = B with 95% confidence level.

These results corroborate that in the second half of April a statistically significant change in the speed of variation of cases occurred in both hemispheres, with an increase in the growth speed of the number of cases in the Southern Hemisphere and a decrease in the growth speed of the number of cases in the Northern Hemisphere. This reinforces the evidence of a consistent, seasonal influence in the transmission of COVID-19.

#### 4. Confirmatory analysis of seasonal effect

We start this section accessing the variability of the seasonal effect. Figure 4 shows the boxplots of the seasonal effects of countries from the Southern and Northern Hemispheres. This emphasizes the difference in the distribution of the effects of the two hemispheres, which corroborates the hypothesis of seasonality.

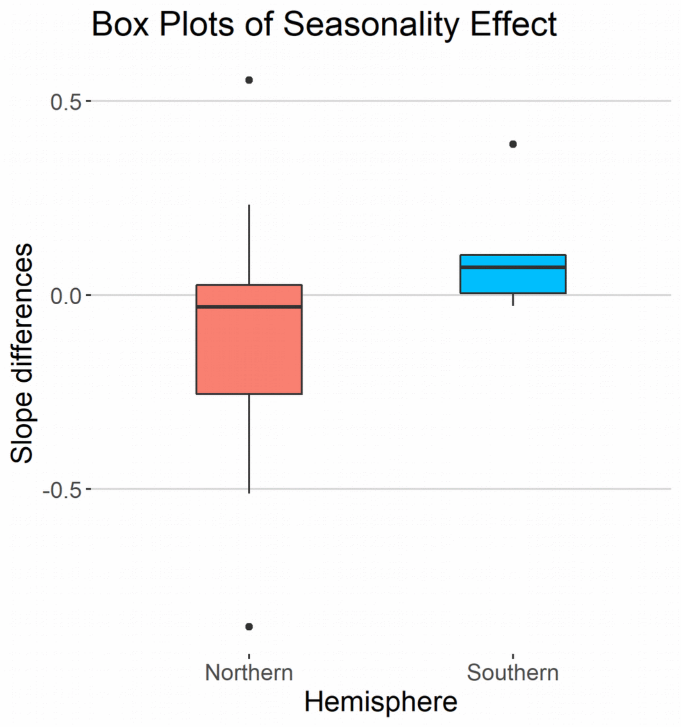


**Figure 4 – Boxplots of seasonal effect:** the boxplot for seasonal effects of countries in the Northern Hemisphere is shown on the left (red boxplot). The boxplot for seasonal effects of countries in the Southern hemisphere is shown on the right (blue boxplot). The majority of the Northern countries had negative slope differences while the majority of Southern countries had positive slopes.

##### 4.1 Methodology validation

As we said before, slope differences for different time intervals naturally occur in epidemic curves. For example, herd immunity is expected to decrease growth speed as the epidemic evolves. Spatial spread in meta-population models, heterogeneous transmission and other factors may also contribute to slope differences. Thus we have to answer how common large slope differences are, like the ones we have obtained in the seasonal effect calculation. To assess the statistical significance of the methodology, we calculated the daily difference between the bounds of confidence intervals for the two slopes and verify other dates when this slope difference was also significant at 95% confidence level.

That is, let [lB,rB] and [lA,rA] be the 95% confidence intervals for slopes B and A defined to calculate the seasonal effect for the mean rate. Where B is the slope of the linear regression line obtained for the mean rate of confirmed cases in the interval [2020-03-27, 2020-04-05], and A is the slope of the linear regression line obtained for data from the interval [2020-04-16, 2020-05-01]. To show that the slope difference is statistically significant, we have to show that either rB < lA in case B < A, or that lB > rA when B>A.

As the slope from the Northern Hemisphere curve presented a decreasing tendency in the period from March to August, we define for each day t, y(t)=lB(t)-uA(t), where A(t) is the slope obtained for the interval [t, t+16] and B(t) is the slope for the interval [t-20, t-11]. For the Southern Hemisphere, we observed a tendency of growth in April; thus, we define for each day t, y(t)=lA(t)-rB(t). We call y(t) the significance difference function at day t.

In both cases, if y(t) > 0, then we have that the slope difference at day t is statistically significant for that hemisphere. Figure S3 shows the significance difference y(t) for both hemispheres from March to August. It shows that the slope differences we used to define the seasonal effect are rarely statistically significant (y>0). The only period when y(t) >0 in both hemispheres is mid-April, which coincides with the estimated onset of the seasonal period.

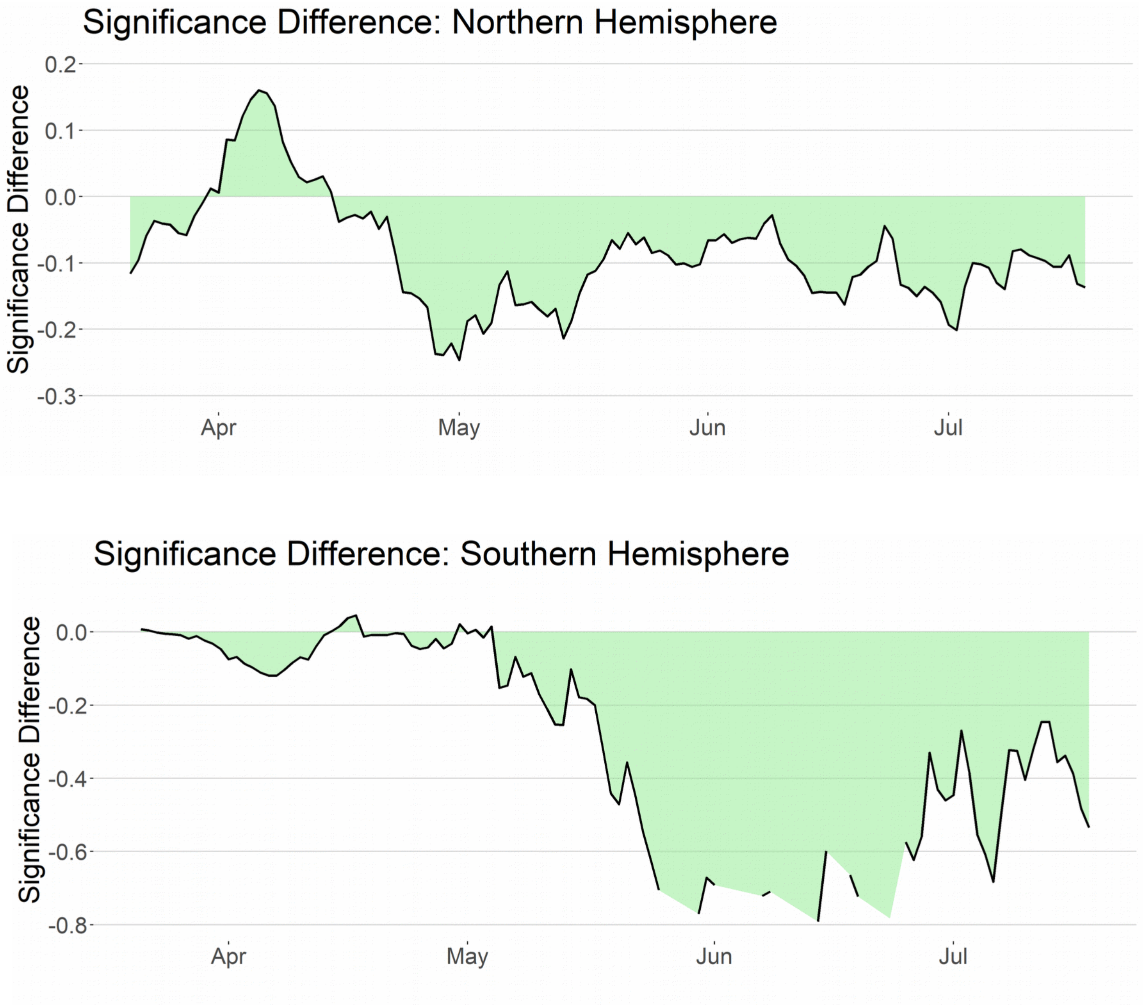


**Figure S3 – Validation of slopes difference method:** Slope differences at day x are statistically significant for y > 0. For both hemispheres, y >0 is uncommon. In the Northern **H**emisphere, y > 0 in the first half of April, and **in** the Southern **H**emisphere y > 0 in mid-April, in some days **at** the end of April and in the beginning of May.

Although slope differences are common in epidemic curves, we conclude that they were only statistically significant in a short period of time, precisely in April. Considering the opposite directions of the slope differences between both hemispheres, this analysis gives us strong statistical evidence that this slope difference in COVID-19 pandemic data is due to seasonal forces, and that its seasonal effect could be detected early on at the end of April.

##### 4.2 Seasonal mean rate time series

The seasonality period can vary from one country to another. Thus, in order to measure seasonality in subgroups of countries, we need a more robust method than the Seasonal effect, which is based on slopes for daily rates of confirmed cases. In addition, when $R0_{min} > 1$, the difference caused by seasonality in the slopes from both hemispheres may not be as appealing as it is when $R0_{min} < 1$, since the rates would keep growing in both hemispheres. Here, we introduce the Seasonal rate that we define to be the daily difference between the mean rate of cases from the Northern Hemisphere and the mean rate of cases from the Southern Hemisphere.

In Figure S4 below, we display the Seasonal rate = mean rate of the Northern Hemisphere – mean rate of the Southern Hemisphere. There are some advantages in measuring seasonal effect through this time series. The seasonal effects from both hemispheres are summed up, which has a number of interesting consequences. The first one is that we have the entire data from both hemispheres integrated in a single curve. The second one is that the seasonal effect is more robust to changes in the time interval used in the regression line after the expected left limit from the seasonal period measured (the seasonal reversal moment). Finally, the third one is that the form of this curve does not depend on whether $R0_{min}$ is greater or lower than one.

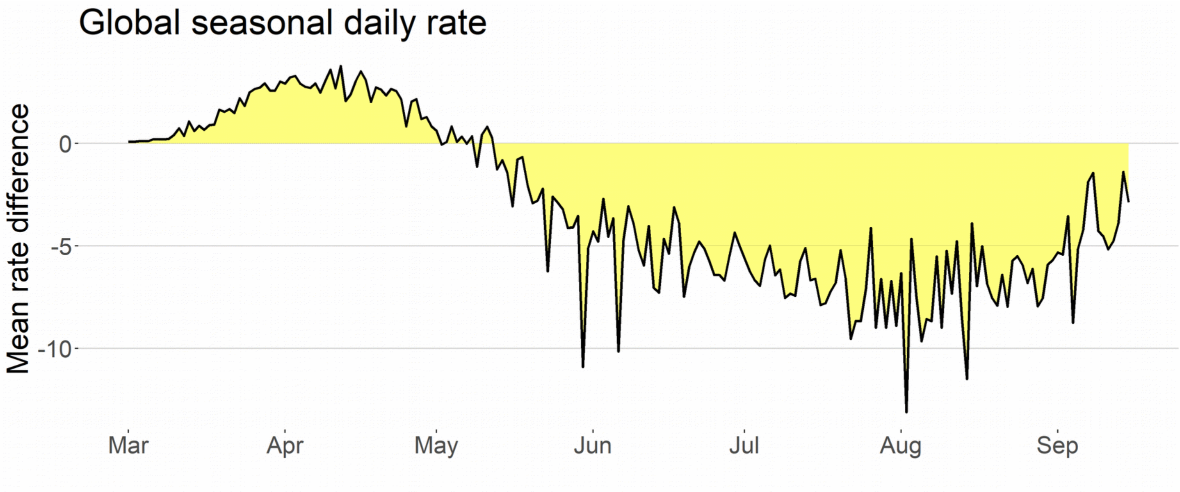


**Figure S4 – Global seasonal rate:** Larger epidemics alternate between the Northern (y>0) and Southern (y<0) Hemispheres. The onset of the high seasons is closely related to the moment when the slope of the seasonal rate curve equals zero. In the COVID-19 pandemic data above, this happened in mid-April. Positive slopes (increasing seasonal rate) are expected in the high season for the Northern Hemisphere and negative slopes (decreasing seasonal rate) are expected in the high season for the Southern Hemisphere.

When the seasonal rate is positive, it means that the mean rate of the Northern Hemisphere is greater than the mean rate of the Southern Hemisphere. On the other hand, when the seasonal rate is negative, we have that the mean rate of cases in the Southern Hemisphere is the greatest. Thus, from Figure S4 we can notice that until May, the Northern Hemisphere was the epicenter of COVID-19 cases, and in the beginning of May, the Southern Hemisphere became the epicenter of the pandemic. Until the end of March, the difference increased in favor of the Northern Hemisphere. In April, there is a turnaround of events and the difference starts to decrease in favor of the Southern cases. This coincides with the beginning of the high season of respiratory seasonal infections in the Southern Hemisphere. From April until August, this tendency of decrease in the seasonal rate remains consistent. A new reversal of the seasonal effect is expected for the beginning of October.

##### 4.3 Subgroup analysis: low-income countries

We repeat the previous analysis, but include only low-income countries in the top 50 epidemics. The inclusion criterion was to have a gross domestic product (GDP) per capita lower than 20,000 dollars. For this analysis, Bolivia is included in the low-income Southern group.

The motivation for this analysis is to verify the hypothesis that the difference in the behavior of the rate of cases between both hemispheres could be explained by economic aspects since, with the exception of Australia and New Zealand, all of the Southern Hemisphere countries are low-income.

We list the countries included below with its respective GDP per capita in dollars in 2018 (source: World bank).

Southern Hemisphere: Argentina (11683), Bolivia (3548), Brazil (8920), Chile (15923), Peru (6941) and South Africa (6374). Mean = 8898 and standard deviation = 4380.

Northern Hemisphere: Bangladesh (1698), Belarus (6289), Dominican Republic (8050), Egypt (2549), India (2009), Iran (5627), Malaysia (11373), Mexico (9673), Panama (15575), Pakistan (1482), Philippines (3102), Poland (15420), Romania (12301), Russia (11288), Serbia (7246), Turkey (9370) and Ukraine (3095). Mean = 7420 and standard deviation = 4724.

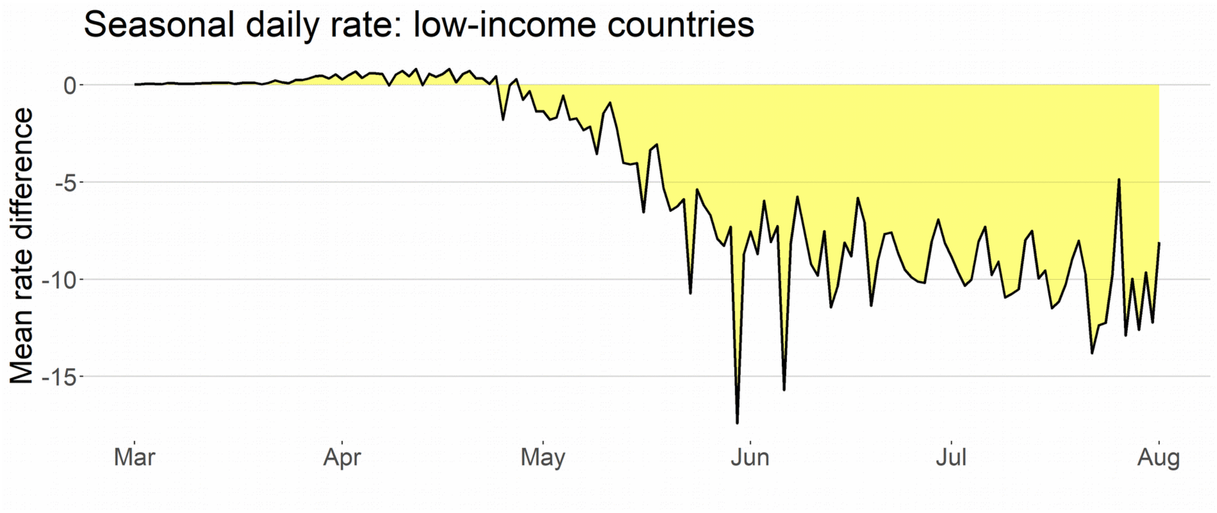

**Figure S5 – Seasonal rate for low income countries:** Larger epidemics alternate between the Northern (y>0) and Southern (y<0) Hemisphere groups. The reversal seasonal moments coincide with the moment when the slope of the seasonal rate curve equals zero. In the COVID-19 pandemic data above, this happened in mid-April. The difference in favor of the group is more pronounced than in the general case of Figure S3.

We observe a qualitative behavior very similar to the previous comparison between all of the countries from both hemisphere groups. The Northern Hemisphere groups present a higher mean rate of cases until April, when we observe a fast change. In May, an inversion occurs, with the average rate of cases in the Southern Hemisphere group presenting more cases. This tendency remains the same during the entire seasonal period that finishes only in September.

Quantitatively, the seasonal differences between low-income countries of both hemispheres are even more pronounced than when mid- and high-income countries are included.

Therefore, this gives data-based evidence that the seasonal effect in the COVID-19 pandemic was even more pronounced in low-income countries. One possible explanation is that the social distancing effect was higher in high-income countries, contributing to a decrease in the rate of cases in high-income countries, regardless of the geographical location. Consequently, we expect a decrease in the seasonal rate, which measures the differences between rates from the Northern and Southern Hemispheres, since both rates would be smaller, softening the natural dynamics of the pandemic. We show in the next subsection that indeed the income factor and the social distancing factor are positively correlated.

##### 4.4 Multiple regression analysis

To corroborate that the difference between the growth speeds from the end of April and the end of March is due to seasonality and not to confounding effects, we run simple and multiple linear regression analyses. We take as response variable Y the seasonality effect (A-B) for each country. The explanatory variables are the seasonal factor $X_{HP}$, the social distancing factor $X_{SD}$ and the income factor $X_{IC}$. We set $X_{HP}$ as the indicator variable if the country belongs to the Northern Hemisphere, $X_{SD}$ as the discrete score varying from 0 to 2 according to the level of social distancing measures adopted (low/none, moderate, high/national lockdown) and $X_{IC}$ as the country's gross domestic product per capita (GDP).

The simple linear regression lines for each explanatory variable are given by $Y = 0.096 - 0.202X_{HP}$, $Y = -0.060 - 0.019X_{SD}$, $Y = 0.004 - 0.094X_{IC}$ and the only statistically significant model at 95% confidence level was the first one for the seasonal factor, with p-value = 0.04. The interpretation for the seasonal factor model is that the mean effect Y = 0.096 for the Southern Hemisphere, which means we have a mean increase of 0.096 for the growth rate of the countries from the Southern Hemisphere. In contrast, the mean effect for countries from the Northern Hemisphere was Y = 0.096 -0.202, that is a reduction of 0.106 in the growth rate.

For the multiple linear analysis, we make a simple re-scaling of the factors $X_{SD}$ and $X_{IC}$.. To do this we just multiply each variable by the average value of $X_{HP}$ and divide them by their respective means. After re-scaling variables to the $X_{HP}$ scale we are able to compare their coefficient estimates and also compare them to the estimates obtained in the simple linear regression for $X_{HP}$. Thus, we obtain for the additive model the estimate Y

= 0.202 -0.184$X_{HP}$ -0.057$X_{SD}$ -0.085$X_{IC}$. All three factors contribute to a reduction on Y, but the seasonal factor $X_{HP}$ has the largest absolute coefficient 0.184 which is similar to the 0.202 obtained in the simple regression.

To assess dependence between factors, we run a multiple linear regression model with interaction terms, obtaining for re-scaled variables the estimate Y = 0.108 -0.162$X_{HP}$ +0.034$X_{SD}$ +0.009$X_{IC}$ -0.011$X_{HP}X_{SD}$ -0.014$X_{HP}X_{IC}$ -0.093$X_{SD}X_{IC}$. We observe that the individual coefficient of the seasonal factor $X_{HP}$ remains the largest, and its estimate is very close to the one obtained in the additive model. The interaction coefficients between seasonal factors and the other two factors are considerably small, which shows that the seasonal factor affected all of the countries, regardless of the levels of social distancing and income factors. The individual coefficients for the social distancing and income factors changed from negative to positive. The reduction measured previously in the additive model is contained in the interaction between the social distancing factor and the income factor. This implies that social distancing effectiveness is highly correlated with income and that its impact was bigger in high-income countries than in low-income ones.

#### 5. Modeling seasonality effect

##### 5.1 Seasonal SEIR model

The seasonal (or time-inhomogeneous) SEIR model is a deterministic model typically used to describe the dynamics of an epidemic in which the transmission rate changes periodically. Suppose we have a population of a fixed size N. The number of people in this population susceptible to the disease at time t is denoted by S(t). The number of people exposed to the infection at time t but not yet capable of transmitting the disease is denoted by E(t). The number of people who are infective at time t is denoted by I(t), and the number of people who are immune to the infection is denoted by R(t). For convenience, we omit time in notation and write simply S, E, I and R, whenever possible. Observe that S + E + I + R = N for all t. Let β(t) denote the rate of transmission at time t, α denote the mean time an infected person is non-infective and γ denote the mean recovery time. The dynamics of the epidemics are given by the following differential equations:

$$\frac{dS}{dt}=-\beta(t)I\frac{S}{N}$$

$$\frac{dE}{dt}=\beta(t)I\frac{S}{N}-\alpha E$$

$$\frac{dI}{dt}=\alpha E-\gamma I$$

$$\frac{dR}{dt}=\gamma I$$

The classical SEIR model is obtained when β(t) is a constant. The above model makes the simplistic assumption that transmission is homogeneously mixed in the population, that is, each infective person transmits the infection at the same rate β(t)S/N. In addition, observe that once an individual is immune to the disease, he remains immune for all t.

The transmission function β(t) can be chosen according to the case of interest. Here, we choose the simplest possible seasonal function, the periodic step function

$$\beta(t)=\beta_{max}, if\ t\in high\ seasonal\ period$$
$$\beta_{min}, if\ t\in low\ seasonal\ period ¿$$

where $\beta_{max} > \beta_{min} > 0$. This function simplifies the analysis of the model, which makes it possible to understand the dynamics in a clear way. Besides its simplicity, it reproduces qualitatively the natural dynamics of seasonal diseases in a short period of a few years. For longer periods, one can use a variation with birth and death rates. Our intention here is to make a qualitative comparison of this model with the classical SEIR in order to understand the effect of seasonality in the qualitative behavior of the dynamics of seasonal diseases. To improve data fitting, a continuous periodic function β(t), such as a trigonometric function, can be chosen or an alternative model, such as the TSIR model, can also be used to model the seasonal effect in epidemiological dynamics (4).

We generate simulated dynamics for the time period from 2020-02-01 to 2021-07-01 for two similar cities, one in the Northern Hemisphere and the other in the Southern Hemisphere. Population sizes are taken to be N = 30000 and initial conditions E=25, I=25 and R=0. We estimate $\alpha = 4.6$ and the mean recovery time $\gamma = 6.75$ (5,*6)*. The high seasonal period is assumed to be from April to September in the Southern Hemisphere, and from September to March in the Northern Hemisphere. The low seasonal periods are the complementary periods of the year.

For the seasonal SEIR with $\beta_t$ transmission function, there are three possible scenarios with different qualitative behavior according to the pair of reproduction numbers ($R0_{min}$, $R0_{max}$). The first case is when $R0_{max} > 1$ and $R0_{min} < 1$. In this case, the epidemics clearly obey the seasonal pattern, with larger outbreaks in the high seasons and a small number of cases in the low seasons. The second case is when $R0_{min} > 1$. Here, one can expect outbreaks starting at any moment of the year, but in the high seasonal period when $\beta = \beta_{max}$, the size and intensity of the outbreaks are larger than in the low seasonal period when $\beta = \beta_{min}$. The last case is when $R0_{max}$ is less than or equal to 1. In this case, there is no tendency that large epidemics will occur at any time, and thus we restrict our analysis to the first two cases and do not show any results on this last situation. For the simulations below, we assume $R0_{max} = 1.5$, $R0_{min} = 0.6$ in the first case and $R0_{max} = 1.5$, $R0_{min} = 1.1$ in the second case. The R0 values take in consideration the fact that social distancing measures reduced the natural transmission rate of COVID-19.

Figures S6 and S7 below show that the dynamics for the Northern and Southern Hemispheres differ considerably from each other in the seasonal SEIR, regardless of whether $R0_{min}$ is bigger or smaller than 1. On the contrary, Figure S7 shows that similar cities will have similar dynamics in both hemispheres in the classical (non-seasonal) SEIR. When $R0_{min} < 1$, epidemic waves alternate from one hemisphere to the other. In this case, the Northern Hemisphere has a small, interrupted first epidemic wave, and a bigger second wave is expected in the next high season. Meanwhile, many cities in the Southern Hemisphere will present a very big first wave and a very small second wave. This resembles the COVID-19 situation of many cities, particularly in Europe.

In case $R0_{min} > 1$ in Figure S6, the situation in the Southern Hemisphere does not change qualitatively. On the contrary, the city in the Northern Hemisphere would have a different dynamic. A continuous, flat first wave is observed during the low season period, until the moment when the high season comes and the number of cases grows faster. However, herd immunity is achieved in a softer manner in comparison to the Southern curve.

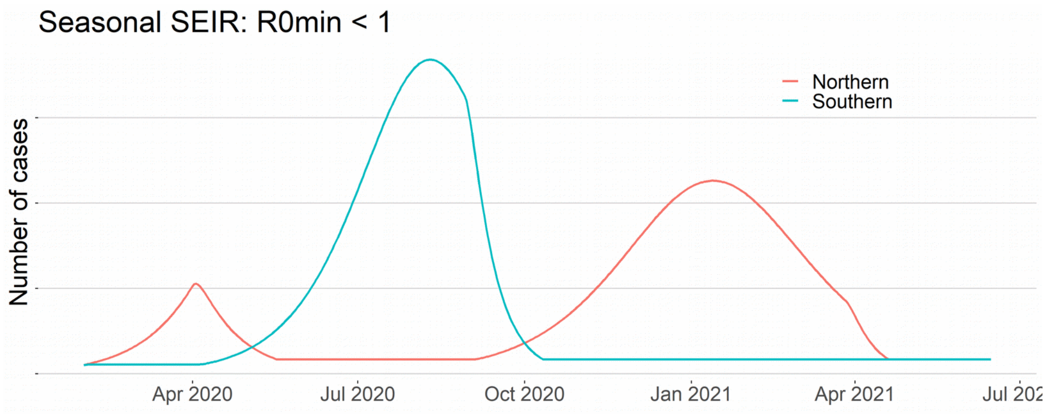

**Figure S6 – Seasonal SEIR: $R0_{min}$ < 1.** Larger epidemics occur in an alternated periodic pattern in the Northern and Southern Hemispheres. Since the initial date was at the end of a Northern seasonal period, we can expect the Northern first wave to be smaller than its second wave (left and right red curves).

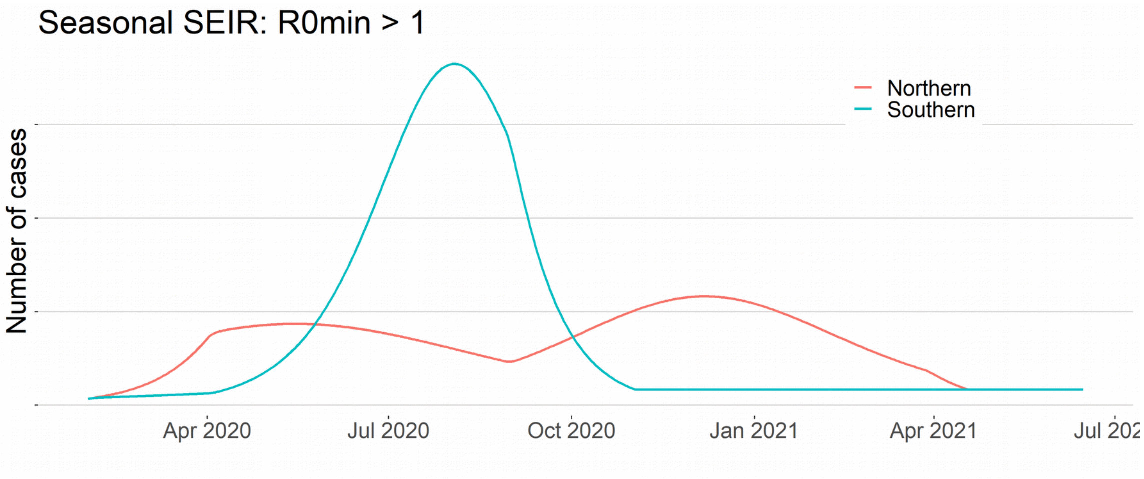


**Figure S7 – Seasonal SEIR: $R0_{min}$ > 1.** Larger epidemics occur in alternated seasonal periods in the Northern and Southern Hemispheres. Here, we observe a long epidemic in the Northern city, with varying growth speed: slower increase in the low season from May to August and a faster increase after September.

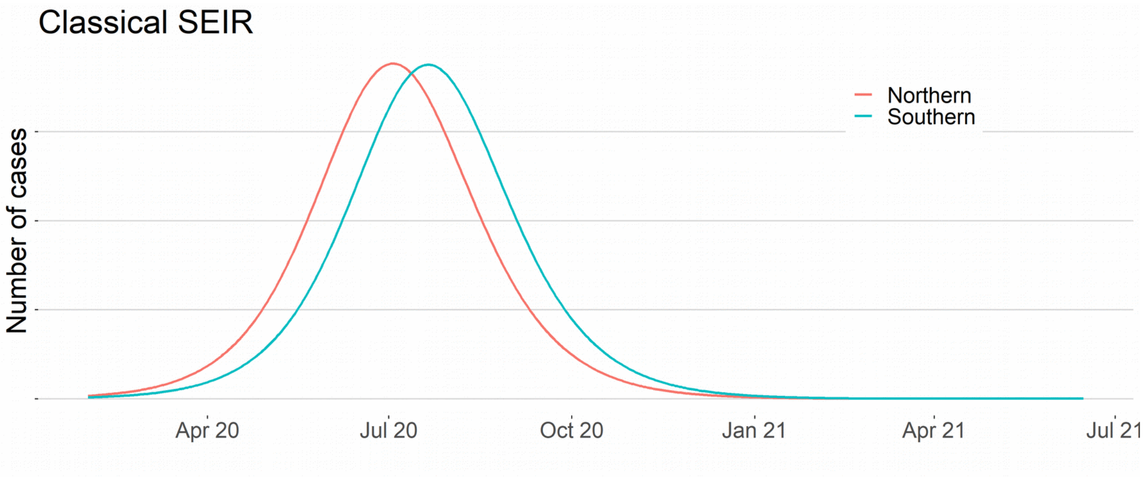


**Figure S8 – Classical SEIR:** Without seasonality, a similar behavior is expected in similar cities **in** the Northern and Southern Hemispheres. Large, single waves occur in similar periods in both hemispheres, and herd immunity controls the moment when the speed of growth changes from positive to negative.

Figure S9 below displays the seasonal rate for simulated seasonal SEIR, when $R0_{min}$ < 1. We observe alternated waves from both hemispheres, with the slopes of the seasonal rate changing signs close to the reversal seasonal moments, when a high seasonal period finishes in one hemisphere and starts in the other (vertical lines). We expect similar dynamics for the COVID-19 seasonal rate.

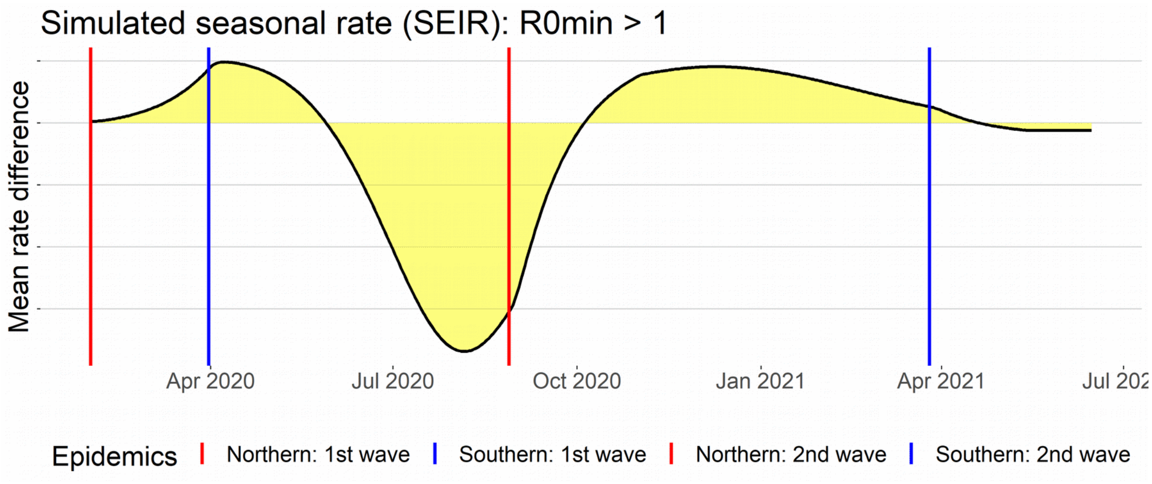


**Figure S9 – Simulated seasonal rate:** Larger epidemics alternate between the Northern (y>0) and Southern (y<0) Hemispheres. The reversal seasonal moments (vertical lines) coincide with the moments when the slope of the seasonal rate curve equals zero, except for a one-week delay. The Northern waves start at the red, vertical lines (positive slopes), and the Southern waves start at the blue, vertical lines (negative slopes). The Northern second wave is bigger than its first wave, which was interrupted by seasonal forces. The Southern first wave is much larger than its second wave, where herd immunity is achieved in a short period.

#### 5.2 Seasonal pattern in COVID-19 data

The epidemic curves in COVID-19 pandemic of many countries are following closely the seasonal pattern of figure S6. To illustrate this we display in figure S10 the mean rate curves for two subgroups of countries from both hemispheres. The Southern group includes Brazil, Peru, Chile, Bolivia and South Africa. Northern group includes the European countries Spain, France, Belgium, Germany, Netherlands, Switzerland, Portugal and the United Kingdom.

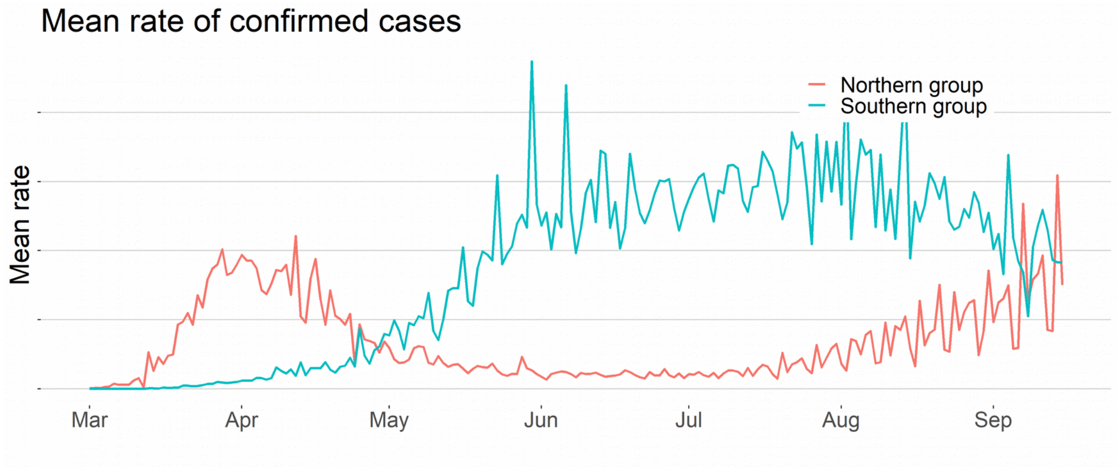

**Figure S10 – Seasonality pattern.** Larger epidemics occur in an alternated periodic pattern in the Northern and Southern Hemispheres. Since the initial date was at the end of a Northern seasonal period, we can expect the Northern first wave to be smaller than its second wave (left and right red curves).

We display below the COVID-19 epidemic curves of daily rates for several countries. High seasonal periods are represented by shaded areas. Countries from the Northern Hemisphere are colored red and countries from the Southern Hemisphere are colored blue. The seasonal pattern of transmission is directly seen in data from these and many other countries.

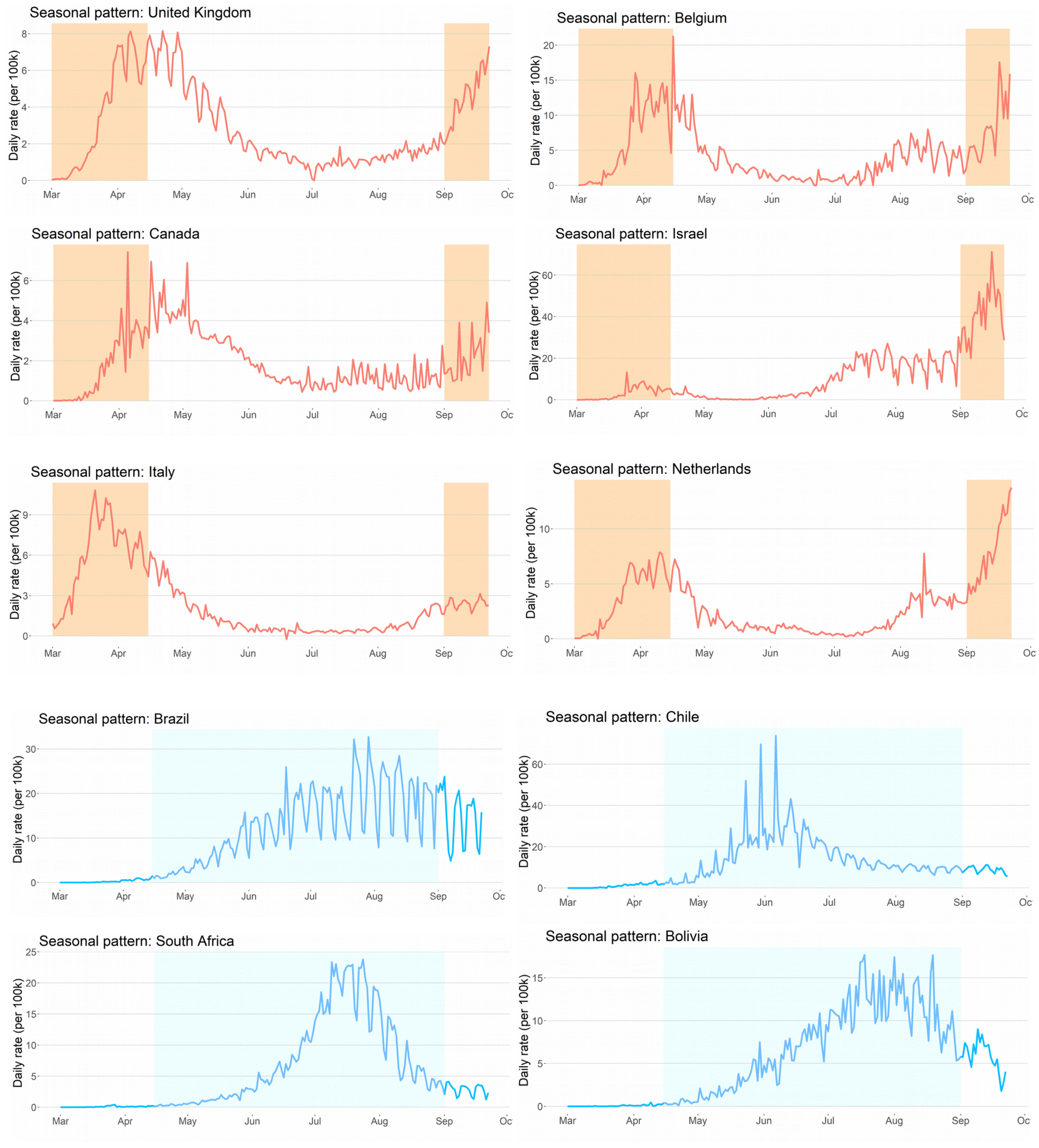

**5.3 Modeling COVID-19 outbreaks**

COVID-19 pandemic was unique in its dynamics, mostly because of the worldwide adoption of social distancing interventions. In this appendix, in section 2, we showed that social distancing interventions had an impact in the growth rate of COVID-19 cases, decreasing its absolute value, causing a flattening of the curves both in Northern and Southern Hemispheres.

In deterministic compartmental models like the SEIR model, this effect can be introduced as a decrease in the transmission rate β(t), as long as the interventions are implemented. Many of the social distancing measures, such as the lockdown, cannot be adopted for long periods. Therefore, in the moment the social distancing interventions are easing, an increase in β(t) is expected.

To study the dynamics of COVID-19 outbreaks, we propose a seasonal SEIR model with an additional parameter δ to represent the social distancing effect. We take δ as a constant number that decreases β(t) during the time interval in which the interventions are adopted. It can be interpreted as the percentage reduction of either the transmission rate β(t) or, equivalently, of the reproduction function R(t). We give priority to parsimony over precision in our choice of a constant effect δ. A decreasing function δ(t) can also be adopted to improve data fitting, as a fatigue effect is expected to occur in long interventions. We adapt the Seasonal SEIR model given in section 5.1 above. The dynamics of the epidemics are given by the differential equations:

$$\frac{dS}{dt}=-(1-\delta(t))\beta(t)I\frac{S}{N}$$
$$\frac{dE}{dt}=(1-\delta(t))\beta(t)I\frac{S}{N}-\alpha E$$
$$\frac{dI}{dt}=\alpha E-\gamma I$$
$$\frac{dR}{dt}=\gamma I$$

As before, we choose the seasonal transmission rate function β(t) as the periodic step function

$$\beta(t)=\beta_{max}, if\ t\in high\ seasonal\ period$$
$$\beta_{min}, if\ t\in low\ seasonal\ period\ ¿$$

where $\beta_{max} > \beta_{min} > 0$. The only difference is given in the social distancing function δ(t) given by

$$\delta(t)=\delta, if\ t\in I$$
$$0, if\ t\notin I\ ¿$$

where I is a union of intervals. Next, in our simulation results, we take $I=[t_1, t_2]$ where $t_1$ is the moment in which the main social distancing interventions started and $t_2$ is the moment in which the main social distancing interventions were eased.

As in section 5.1, we generate a simulated dynamics for the time period from 2020-02-01 to 2021-07-01 for two similar cities, one in the Northern Hemisphere and the other in the Southern Hemisphere. Population sizes are taken to be N = 25000 and initial conditions E=10, I=10 and R=0. We estimate $\alpha = 4.6$ and the mean recovery time $\gamma = 6.75$ (5,*6)*. The high seasonal period is assumed to be from April 15 to August 31in the Southern Hemisphere and from September 01 to April 14 in the Northern Hemisphere. The low seasonal periods are the complementary periods of the year.

As in section 5.1, for the seasonal SEIR with $\beta(t)$ transmission function, there are three possible scenarios with different qualitative behavior according to the pair of reproduction numbers ($R0_{min}$, $R0_{max}$). The first case is when $R0_{max} > 1$ and $R0_{min} < 1$. In this case, the epidemics obey the seasonal pattern clearly, with larger outbreaks in the high seasons and small number of cases in the low seasons. The second case, when $R0_{min} > 1$, is the most complex. Here, one can expect outbreaks starting at any moment of the year, but in the high seasonal period when $\beta = \beta_{max}$, the size and intensity of the outbreaks are larger than in the low seasonal period when $\beta = \beta_{min}$ . The last case is when $R0_{max}$ is less than or equal to 1. In this case, there is no tendency to occur large epidemics at any time, thus we restrict our analysis to the first two cases and do not show any results on this last situation. For the simulations below, we assume $R0_{max} = 2.0$, $R0_{min} = 0.9$ in the first case and $R0_{max} = 2.0$, $R0_{min} = 1.4$ in the second case. We expect that, for COVID-19, in many cities R0min > 1.

We set the social distancing constant $\delta = 35\%$ or 70%, representing less effective or more effective interventions, respectively. In figure S11, for R0min < 1, seasonality drives the outbreaks. Effective social distancing interventions can decrease epidemic size or delay it, but if no other control measures are used, a bigger second wave is expected.

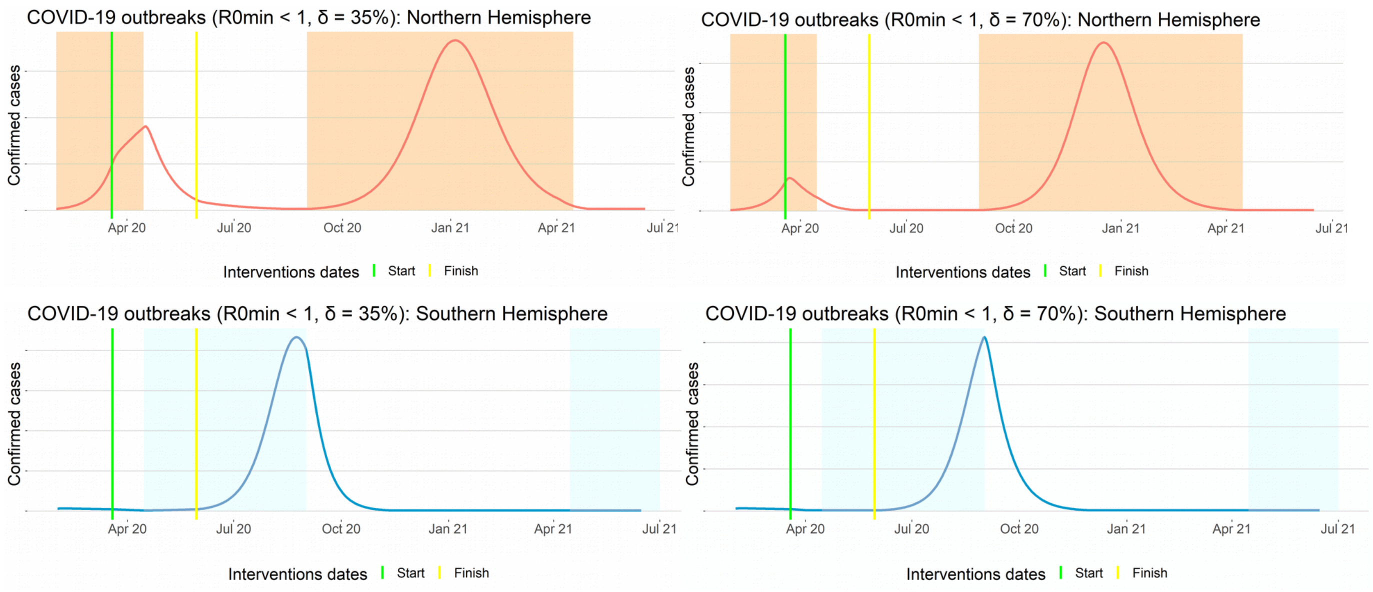


**Figure S11 – COVID-19 compartmental model: R0min < 1.** Seasonality drives the outbreaks patterns in all cases if R0min < 1. Green vertical lines show the start intervention date and yellow vertical lines show the date when social distancing interventions were eased. Effective social distancing effect ($\delta = 70\%$) decreased the first wave in the Northern Hemisphere, but after relaxation, produced a larger second wave in the next high seasonal period.

In figure S12, for R0min > 1 and effective social interventions (δ = 70%), a similar dynamics is observed. Seasonality also drives the outbreaks, but effective social distancing interventions can decrease epidemic size or delay it. However, if no other control measures are used such as vaccination or effective contact tracing, once social distancing interventions are relaxed, a bigger second wave is expected.

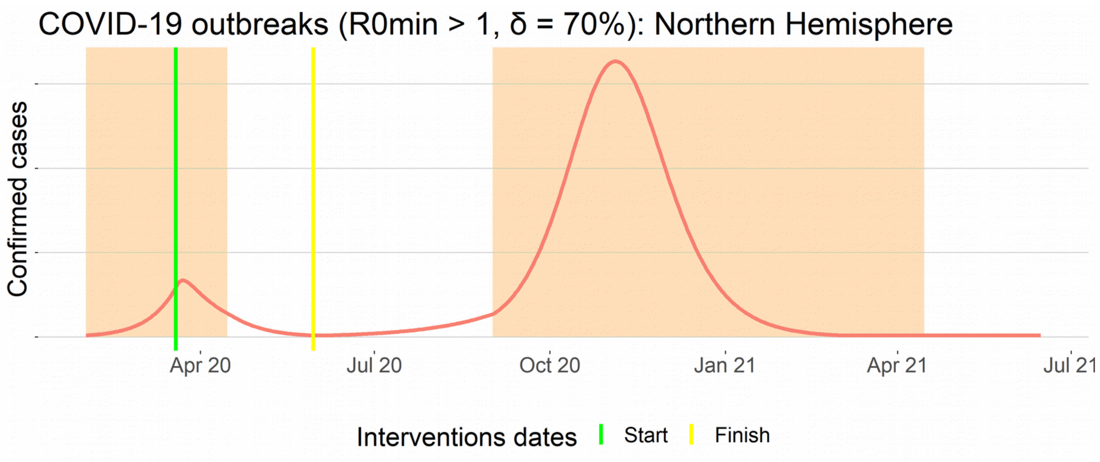


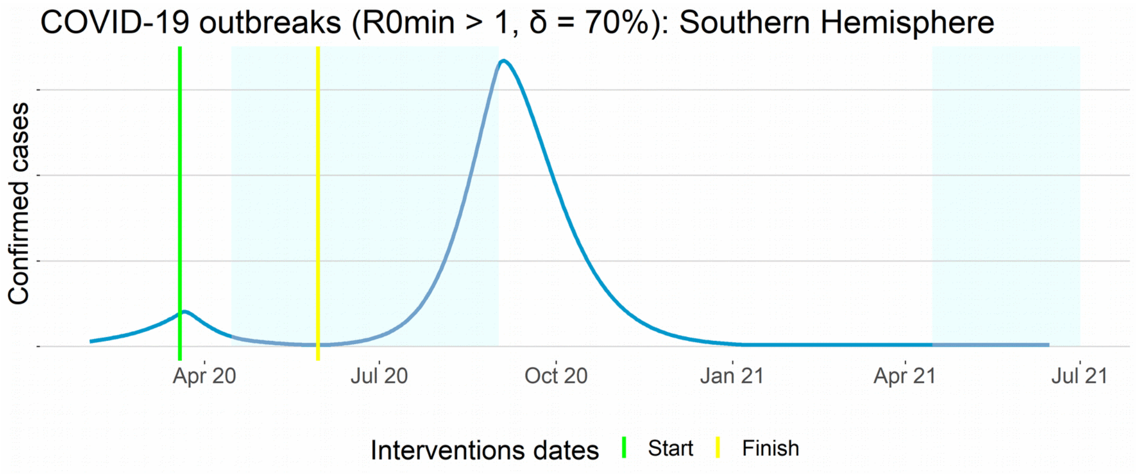


**Figure S12 – A compartmental model for COVID-19: R0min > 1 and δ = 70%.** Outbreaks follow seasonal patterns closely, if the social distancing effect is large. Social distancing interventions decreased the size of first wave in the Northern Hemisphere, but once relaxed, it produces a larger second wave in the next high seasonal period, starting in September. In the Southern Hemisphere, an effective social distancing intervention can control the epidemics even in the high seasonal period.

For R0min > 1 and less effective social interventions (δ = 35%), we observe more complex patterns arising. In figure S13 below, in the Northern Hemisphere, social distancing effect flattened the curve (green line), but did not control the epidemics. When seasonal forces reduce transmission rate (white rectangle), the number of cases starts to decrease, but once the interventions are relaxed (yellow line) the number of cases increases again. In September, in the onset of the high seasonal period (red shaded rectangle), transmission rate increases and the outbreak accelerates. A similar pattern can be seen in real world COVID-19 data, for example in the epidemic curve of the United States (see Figure S14). For the Southern Hemisphere, social distancing interventions started at the end of the low seasonal period (green line). It also flattened the curve, but in the onset of the high seasonal period (blue shaded rectangle) the number of cases rose, in spite of the social distancing measures, although we note that the growth rate is reduced. When social distancing effect is lost in the middle of the high season (yellow line), the transmission speeds up until herd immunity is achieved and cases start to decrease. When the low season comes (left white rectangle), the number of cases decreases faster until the end of the epidemics.

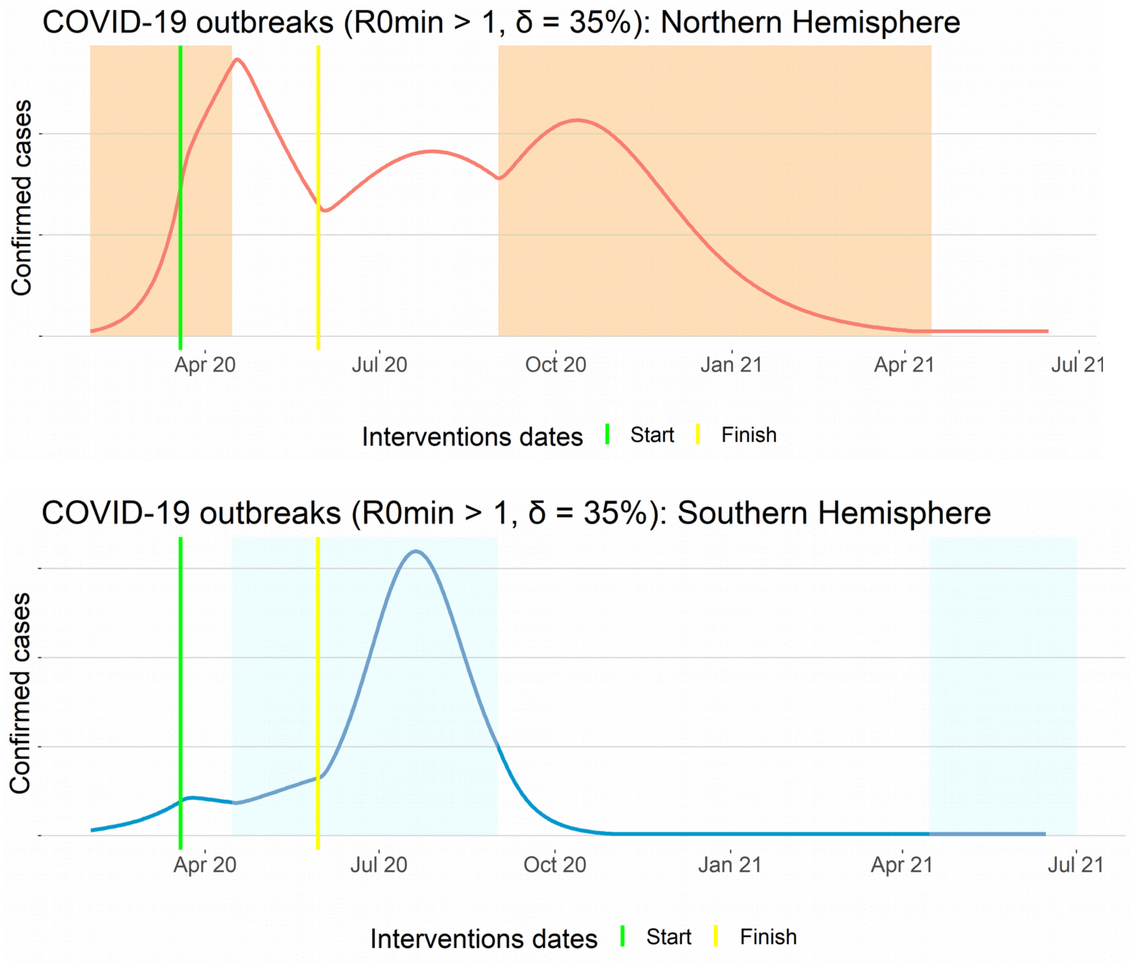


**Figure S13 – A compartmental model for COVID-19: R0min > 1 and δ = 35%.** Outbreaks follow seasonal patterns rigidly in all cases., when R0min < 1. Larger social distancing effect (δ = 70%) decreased the first wave in the Northern Hemisphere, but after relaxation, produced a larger second wave in the next high seasonal period. Green vertical lines show the start intervention date and yellow vertical lines show the date when social distancing interventions were eased.

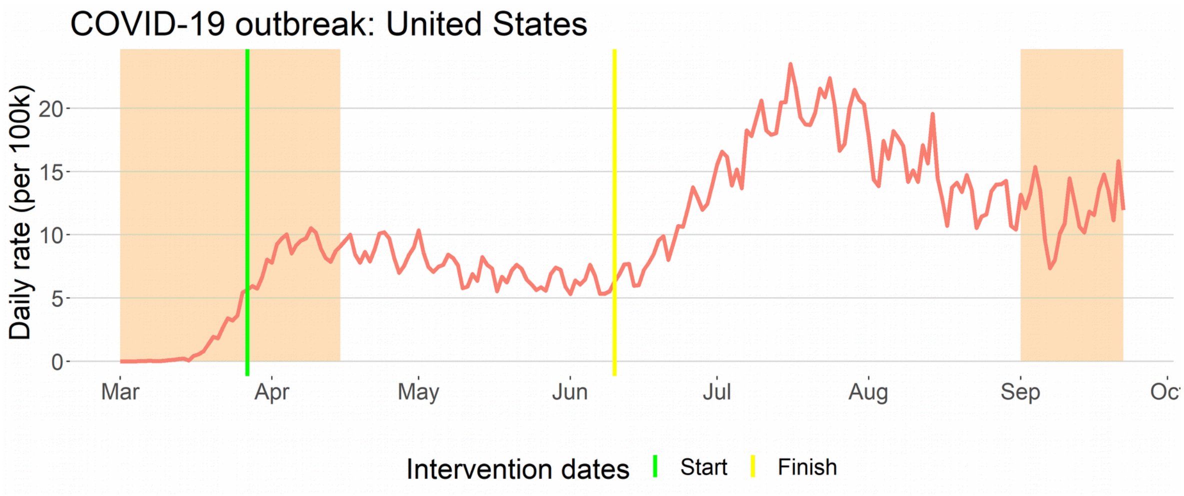


**Figure S14 – Seasonal pattern in COVID-19 outbreak in the United States.** United States reported data is an example of the complex pattern obtained when R0min > 1 and social distancing interventions affect transmission rate, just like in Figure S12. Growth rates change when the seasonal period changes in mid-April and in September due to seasonal forces. Between green and yellow vertical lines social distancing effect decreases the transmission rate. Both social distancing effect and seasonal effect are fundamental to explain the outbreak dynamics. Note: the yellow line was defined as the mean date of the reopening of gyms, bars and restaurants in the states of California, Texas and Florida, which concentrated the majority of the cases in this period of time.

### 6. Limitations

As a database, reported cases have many limitations. There is a significant amount of unreported cases and a large variability in the quality and reliability of the data from country to country. As we used polled data methods to analyze the data, we have minimized the variability problem. In addition, we focused on data from the larger reported epidemics. There are many countries with weak developed epidemiological surveillance, with very small reported epidemics. By including only the largest epidemics, we avoid the inclusion of this less reliable data.

However, there were still some particular limitations which deserved some attention. In this section, we examine two of these limitations.

#### 6.1 Reported cases in China

The analyses of this article rely on a time-series perspective. We used data from daily reported confirmed cases; thus, it is necessary that each country follow a daily report rule. We have chosen to restrict our pool to the top 50 countries with the largest number of confirmed cases from 2020-03-01 to 2020-05-01. A possible more natural alternative is to restrict to a period starting from the beginning of the pandemic until May 1. The difference between the two obtained groups is merely the inclusion of China in the last ranking, instead of Finland.

However, the confirmed cases in China did not follow a daily basis report. A significant part of cases was reported in two days, as we can see in Figure S15 below. This problem also occurred in other countries, but for China this happened coincidentally in the moment in which we measured the seasonal effect. Second, China was the country where the pandemic started, probably earlier than January 2020. It has adopted an aggressive lockdown even before the epidemics reached many of the countries in the pool, which makes its dynamics not synchronized with the rest of the world. For this reason, we have decided to exclude it from the pool and include Finland instead.

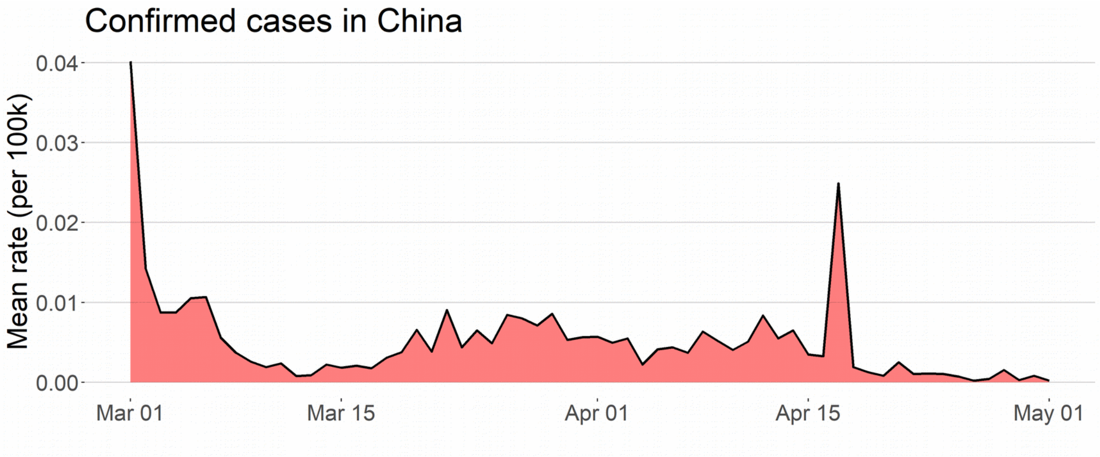


**Figure S15 – Daily rate of confirmed cases of COVID-19 in China:** A single day concentrates many of the reported cases.

#### 6.2 Regional seasonal patterns

In our analysis we associated seasonality and measured its effects subdividing the countries into two main groups, Southern and Northern Hemisphere groups. Formally, we ran our analysis using the indicator random variable which attributes the value 1 to countries in the Northern Hemisphere and 0 to countries in the Southern Hemisphere. As previously said, seasonality is a complex phenomenon and the use of any single factor will end up with some level of error. For example, choosing temperature as a factor to associate to seasonality implies in relative errors, since a temperature of 15 Celsius degrees can be either summer in one country or winter in another.

In our case, using the division Northern and Southern Hemispheres disregards some well-known regional seasonal patterns. Here we give one important example: Latin American countries. Although Mexico and Central America countries are in the Northern Hemisphere, for seasonal respiratory viruses, they follow a similar pattern, which coincides with the Southern Hemisphere seasonal pattern (7). Thus, Mexico, Panama and Dominican Republic, despite being allocated in the Northern Hemisphere group, follow closely the seasonal pattern from the Southern Hemisphere (see figure S12, below).

As a consequence, this introduces an error in the analysis. However, note that, on average, this error decreases both seasonal effects and statistical significance of the tests used. Therefore, changing these countries from the Northern Hemisphere group to the Southern Hemisphere group would cause a positive effect on our results.

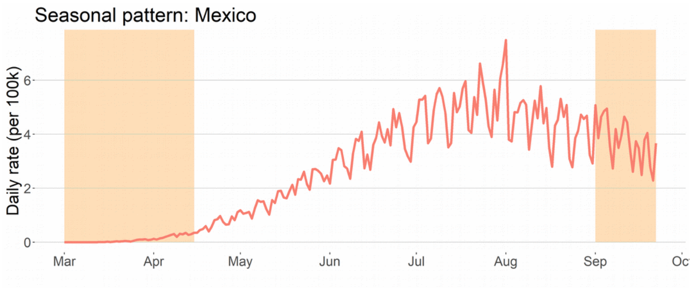


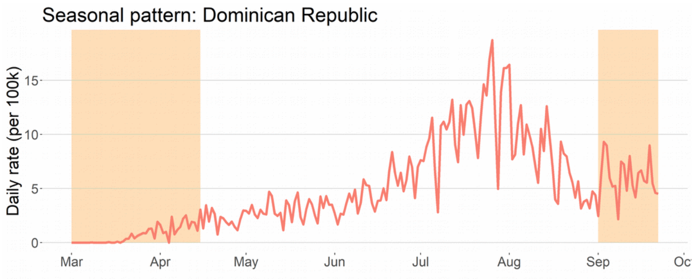


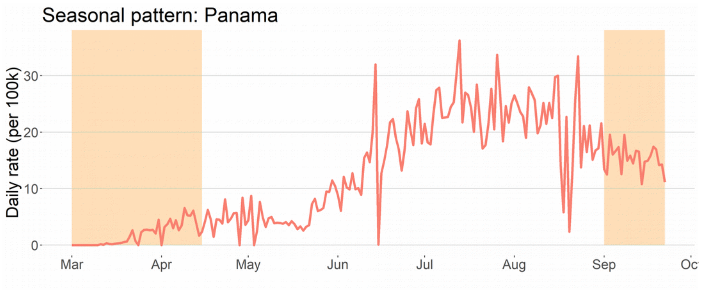

**Figure S16 – Latin America seasonal patter:** Countries from Latin America, including Northern Hemisphere countries such as Mexico, Panama and Dominican Republic, follow the seasonal pattern from the Southern Hemisphere with a higher transmission rate from April to August.

#### 6.3 Case definitions

The number of confirmed cases is the main information released concerning the dynamics of the COVID-19 pandemic in each country. This fundamental epidemiological surveillance data is used by governments, scientists and society to make projections and measure the burden of the disease.

In most countries, the definition of a confirmed case follows World Health Organization-WHO guidance. The official definition states that a person becomes a confirmed case only in the presence of a laboratory confirmation (*8*). A recent study shows that different definitions can significantly impact important epidemiological variables (9).

The crucial issue here is that laboratory tests, such as the molecular RT-PCR test, are intended to detect the specific virus with high precision. On the other hand, serology tests are imprecise and are not included in this definition, as recommended by the WHO itself (*10*). A significant number of serology tests produce false positive results, typically in the range of 0.7% to 7.5% (*11*).

In addition, there is also a high rate of false negatives. In order to measure the possible impact this error could cause, note the Brazilian Ministry of Health has requested more than 20 million serology test kits, which implies that the number of confirmed cases could increase by up to 1.5 million due to false positives (*12*). Until June 23, Brazil officially had 1,145,906 confirmed cases, which represents about 0.5% of the total population.

Cases with a clinical diagnosis or with only a positive serology test are considered only probable cases by the WHO (8). The European Union, the United States, Canada, Argentina, Chile, South Korea and Japan are some of the many countries that follow these definitions (13-*19*).

Brazil has its own definition of a confirmed case, which includes laboratory confirmed cases, clinical suspicious cases that had contact with a confirmed case and cases that tested positive in serology tests (*20*). According to the WHO, these last two should be considered probable cases. Until May 9, the proportion of confirmed cases that had a test confirmation by molecular RT-PCR was 67.6%, while the proportion of cases confirmed by serology tests was 32.4%. In addition, 1.4% of all of the confirmed cases were clinical cases (*21*). On May 29, the proportion of cases confirmed by RT-PCR reduced to only 56% (*22*). Thus, up to this date, almost one in two of the tested confirmed cases in Brazil should not be considered a confirmed case (10,*23*).

To access the impact of serology tests on reported cases, we fix the rate of clinically confirmed cases at 1.4%. Since May 9, we estimate that 47.3% of confirmed cases were confirmed through serology tests. Thus, 48.7% of the new cases should be classified as probable cases. We estimate that 51.3% of the cases reported in Brazil after May 9 are laboratory confirmed cases. This estimate agrees with the proportion of 54.3% of molecular tests in Brazil, which has requested 22 million serology tests and 24.6 million molecular RT-PCR tests (*12*). The number of officially confirmed cases on May 9 was 155,939 and on May 29 was 465,166. Therefore, the corrected number of confirmed cases in Brazil can be estimated by the formula:

$$Estimated\ confirmed\ cases = 103,939 + 0.513 \times (officialy\ confirmed\ cases - 155,939)$$

On June 23, the number of confirmed cases was 1,145,906. By applying the formula above, we obtain the estimate of 611,792 for the corrected number of confirmed cases and 534,114 for the number of probable cases.

In Brazil, molecular RT-PCR tests are mainly used in hospitalized patients and deaths. Thus, we consider the number of officially confirmed deaths as an approximation to the estimated number of confirmed deaths. We estimate the corrected lethality rate by the formula:

$$Corrected\ lethality\ rate = \frac{(officially\ confirmed\ deaths)}{(Estimated\ confirmed\ cases)}$$

On June 23, the number of confirmed deaths was 52,645. By applying the formula above, we obtain the corrected lethality rate of 8.6%, in contrast to the official rate of 4.6%, which implies a relative increase of 87.5%. Hence, caution is necessary when comparing the reported cases from Brazil with the reported cases from other countries. To make our main analysis, we only used data from March and April. Thus, the impact of Brazil's definition of cases is limited, as the serology tests were more widely used after May. Nevertheless, our main measure to assess seasonality is the mean rate, which gives Brazil the same weight as to any other country. This also contributes to limiting a possible negative impact in our analysis.

Besides Brazil, the United States also deviates from the standards when disclosing its results. Despite following WHO definitions, the United States Centers of Disease Control and Prevention (CDC) discloses the number of total cases, which includes confirmed cases plus probable cases (8,*14*,*15*). However, the percentage of laboratory confirmed cases in the United States corresponded to around 96.6% of the total cases reported on June 26 (*24*). Therefore, the impact of probable cases in the total reported cases in the United States' data in March and April is considerably small.

Corresponding author: Marcio Watanabe


Instituto de Matemática e Estatística,
Universidade Federal Fluminense, Brazil.

[10] World Health Organization.
Advice on the use of point-of-care immunodiagnostic tests for Covid-19.

https://www.who.int/news-room/commentaries/detail/advice-on-the-use-of-point-of-care-immunodiagnostic-tests-for-covid-19
Date: 2020.
Date accessed: May 25, 2020.

[11] Coronavirus kit evaluation program (in Portuguese)
https://testecovid19.org/avaliacoes/
Date: 2020.
Date accessed: May 25, 2020.

[12] Brazilian Ministry of Health.
https://www.saude.gov.br/noticias/agencia-saude/46760-ministerio-da-saude-amplia-para-46-2-milhoes-aquisicao-de-testes
Date: 2020.
Date accessed: May 25, 2020.

[13] European Centre for Disease Prevention and Control.
Case definition for coronavirus disease.
https://www.ecdc.europa.eu/en/case-definition-and-european-surveillance-human-infection-novel-coronavirus-2019-ncov
Date: 2020.
Date accessed: May 25, 2020.

[14] United States Centers for Disease Control and Prevention.
Coronavirus disease 2019.
https://wwwn.cdc.gov/nndss/conditions/coronavirus-disease-2019-covid-19/case-definition/2020/
Date: 2020.
Date accessed: May 25, 2020.

[15] United States Centers for Disease Control and Prevention.
Coronavirus disease 2019: cases in the U.S.
https://www.cdc.gov/coronavirus/2019-ncov/cases-updates/cases-in-us.html
Date: 2020.
Date accessed: May 25, 2020.

[16] Canada government.
Interim national case definition.
https://www.canada.ca/en/public-health/services/diseases/2019-novel-coronavirus-infection/health-professionals/national-case-definition.html
Date: 2020.
Date accessed: May 25, 2020.

[17] South Korea government.
Case definition.

http://ncov.mohw.go.kr/en/baroView.do?brdId=11&brdGubun=112&dataGubun=&ncvContSeq=&contSeq=&board_id=&gubun=
Date: 2020.
Date accessed: May 25, 2020.

[18] Argentine government.
Case definition.
https://www.argentina.gob.ar/salud/coronavirus-COVID-19/definicion-de-caso
Date: 2020.
Date accessed: May 25, 2020.

[19] Chile government.
Case definition.
https://www.minsal.cl/wp-content/uploads/2020/04/Ord.-B51-N%C2%BA933.pdf
Date: 2020.
Date accessed: May 25, 2020.

[20] Brazilian Ministry of Health.
Case definition and notification.
https://coronavirus.saude.gov.br/definicao-de-caso-e-notificacao
Date: 2020.
Date accessed: May 25, 2020.

[21] Brazilian Ministry of Health.
Epidemiological bulletin 16.
https://portalarquivos.saude.gov.br/images/pdf/2020/May/21/2020-05-19---BEE16---Boletim-do-COE-13h.pdf
Date: 2020.
Date accessed: May 25, 2020.

[22] CNN Brazil website: data from Brazilian Ministry of Health (in Portuguese).
https://www.cnnbrasil.com.br/saude/2020/06/04/ate-maio-27-8-testes-rt-pcr-para-covid-19-deram-positivo
Date: 2020.
Date accessed: June 04, 2020.

[23] United States Centers for Disease Control and Prevention.
Evaluating and testing persons for coronavirus disease.
https://www.cdc.gov/coronavirus/2019-ncov/hcp/clinical-criteria.html
Date: 2020.
Date accessed: May 25, 2020.

[24] United States Centers for Disease Control and Prevention.
CDC COVID data tracker.
https://www.cdc.gov/covid-data-tracker/index.html#cases
Date: 2020.
Date accessed: June 16, 2020
